# Human Cytomegalovirus mRNA-1647 Vaccine Candidate Elicits Potent and Broad Neutralization and Higher Antibody-Dependent Cellular Cytotoxicity Responses than that of the Partially Effective gB/MF59 Vaccine

**DOI:** 10.1101/2023.08.02.23293386

**Authors:** Xintao Hu, Krithika P. Karthigeyan, Savannah Herbek, Sarah M. Valencia, Jennifer A. Jenks, Helen Webster, Itzayana G. Miller, Megan Connors, Justin Pollara, Caroline Andy, Linda M. Gerber, Emmanuel B. Walter, Kathryn M. Edwards, David I. Bernstein, Jacob Hou, Matthew Koch, Lori Panther, Andrea Carfi, Kai Wu, Sallie R. Permar

## Abstract

**Background:** The MF59-adjuvanted gB subunit (gB/MF59) vaccine demonstrated ~50% efficacy against human cytomegalovirus (HCMV) acquisition in multiple clinical trials, suggesting efforts to improve this vaccine design might yield a vaccine suitable for licensure. A vaccine candidate employing nucleoside-modified mRNAs encoding HCMV gB and pentameric complex (PC) encapsulated in lipid nanoparticle, mRNA-1647, is currently in late-stage efficacy trials. Yet, its immunogenicity has not been compared to the partially-effective gB/MF59 vaccine.

**Methods:** We assessed neutralizing and Fc-mediated IgG effector antibody responses induced by mRNA-1647, a vaccine comprising an equal mass of 6 mRNAs encoding gB and PC antigens, in both HCMV seropositive and seronegative vaccinees from a first-in-human clinical trial through 1-year following 3^rd^ vaccination using a systems serology approach. Further, we compared peak anti-gB antibody responses in seronegative mRNA-1647 vaccinees to that of seronegative female adolescent gB/MF59 vaccine recipients.

**Results:** mRNA-1647 vaccination boosted pre-existing HCMV-specific IgG responses in seropositive vaccinees, including neutralizing and Fc-mediated effector antibody responses. In seronegative vaccinees, mRNA-1647 induced durable and functional HCMV-specific IgG responses. Elicited gB-specific IgG responses were lower than the PC-specific IgG responses. Additionally, gB-specific IgG and antibody-dependent cellular phagocytosis (ADCP) responses were lower than those elicited by gB/MF59. However, mRNA-1647 elicited robust neutralization and high antibody-dependent cellular cytotoxicity (ADCC) responses.

**Conclusions:** mRNA-1647 vaccination induced polyfunctional and durable HCMV-specific antibody responses. mRNA-1647-elicited gB-specific IgG responses were lower than PC-specific IgG responses and lower than those elicited by the partially effective gB/MF59. However, higher neutralization and ADCC responses were elicited by mRNA-1647 than gB/MF59.

**Clinical Trials Registration:** ClinicalTrials.gov (NCT03382405, mRNA-1647) and (NCT00133497, gB/MF59).

**Summary:** mRNA-1647 HCMV vaccine elicited polyfunctional and durable antibody responses in humans. While the mRNA-1647-elicited glycoprotein B (gB)-specific IgG responses were lower than that of the moderately-effective gB/MF59 vaccine, the pentameric complex (PC)-specific IgG responses were strong.

## INTRODUCTION

Human cytomegalovirus (HCMV), a ubiquitous herpes virus, leads to life-threatening diseases in newborns infected in utero (1) and in immunocompromised individuals (2), with severe symptoms such as enteritis, pneumonitis, and neurologic deficits (1, 2), highlighting the need for an effective vaccine to prevent infection and disease (3, 4). Viral attachment is mediated by major envelope glycoprotein complexes on the virion surface (3, 5), including glycoprotein B (gB), the primary viral fusion protein (6), and gH/gL/UL128/UL130/UL131A pentametric complex (PC), which is essential for entry into epithelial, endothelial, dendritic, and monocytic cells (7). Both gB and PC are viable targets for antibody based anti-CMV interventions (3, 8).

MF59-adjuvanted HCMV Towne gB subunit vaccine (gB/MF59) is the most efficacious HCMV vaccine tested to date, with approximately 50% protection in several phase 2 trials (9–12). Yet, efficacy was not high enough to advance to late-stage clinical trials. Therefore, efforts are needed to develop a better vaccine that elicits more protective responses. Interestingly, this vaccine achieved moderate protection with only limited neutralizing antibody (NAb) responses, suggesting a role for non-NAb responses in preventing HCMV acquisition (9, 13, 14). However, inducing broad and potent NAb responses might also enhance vaccine efficacy (3).

Frequent reinfection and reactivation in HCMV-exposed individuals represent another challenge to preventing and controlling disease (15). Vaccination may enhance immunity to prevent reinfection or end-organ disease in affected populations (3). Although the precise immune correlates of protection against HCMV acquisition, congenital HCMV (cCMV) transmission, or end-organ disease have yet to be established, there is growing evidence that Fc-mediated effector antibody functions are important in preventing cCMV transmission (16, 17). Adding antigens that induce potent NAb and Fc-mediated effector responses might elicit polyfunctional humoral responses that improve vaccine efficacy (3, 8).

The modified mRNA encapsulated in lipid nanoparticle (LNP) vaccine platform has remarkable advantages for both manufacturing ease and safe induction of effective immunity in humans (3, 18). Toward this end, Moderna developed an HCMV vaccine candidate, mRNA-1647, comprised of both gB and PC (19, 20), which induced durable and functional antibody responses in preclinical studies in mice and non-human primates (NHP) (19, 20). In this report, we comprehensively assessed the polyfunctional humoral responses elicited by mRNA-1647 in HCMV seronegative and seropositive vaccinees and compare the gB-specific IgG responses with those induced by the partially effective gB/MF59 vaccine.

## METHODS

### Study Participants and Design

Information about study participants is summarized in Table 1. Both gB/MF59 and mRNA-1647 vaccines were delivered via intramuscular injection (IM) in a three-dose series at months 0, 1 or 2 and 6 (Figure 1A). gB/MF59 samples came from a Phase 2 study in healthy adolescent females (NCT00133497). mRNA-1647 samples are from a Phase 1 study in healthy adults (NCT03382405). Information about study products and design is detailed in supplementary materials.

**Figure 1.**
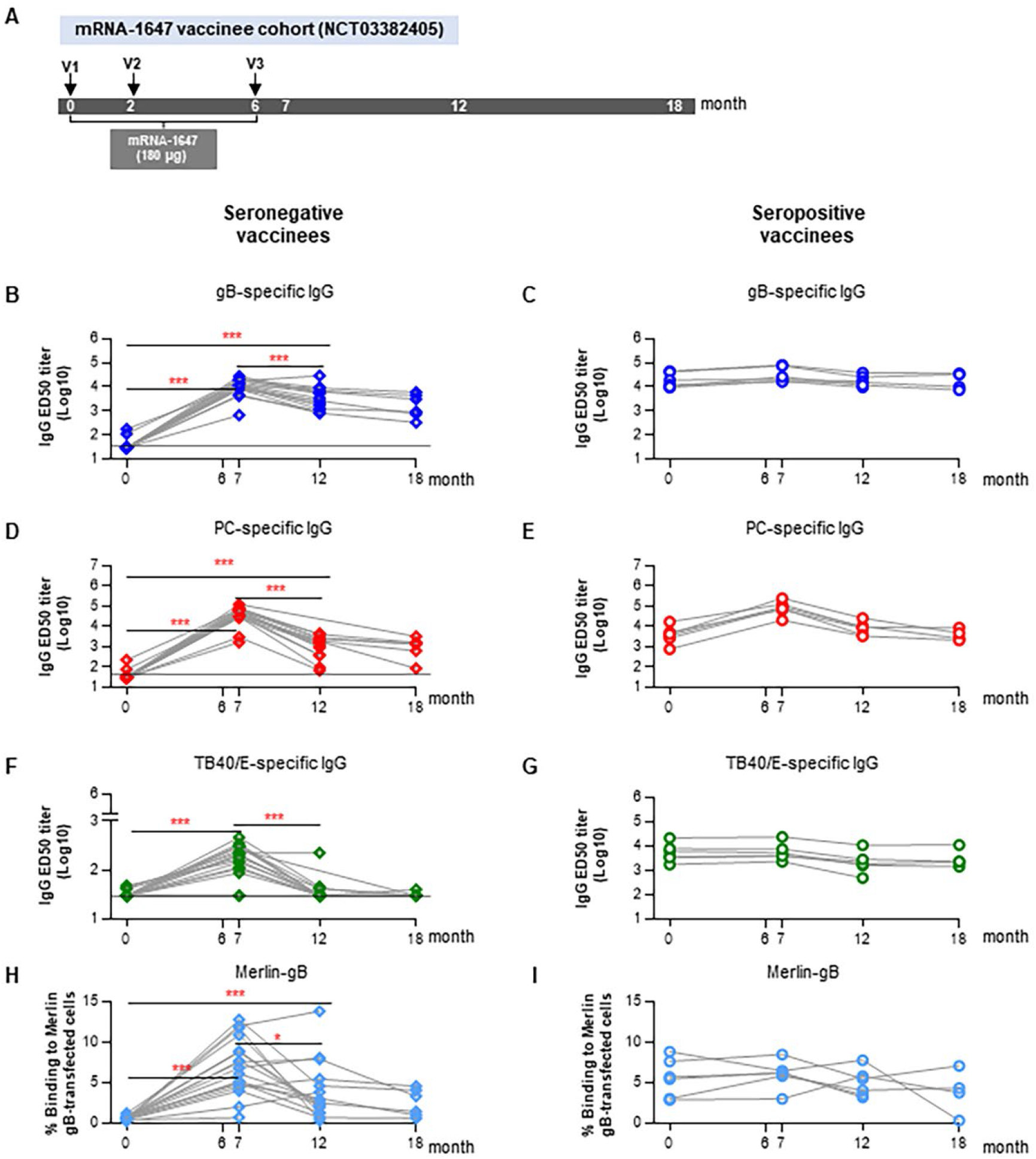
mRNA-1647 vaccine-induced long-lasting antigen-specific IgG antibody responses in HCMV seropositive and seronegative vaccinees. (A) The immunization schedule for HCMV mRNA-1647 vaccine candidate with doses administrated at months 0, 2, and 6 for both HCMV seropositive (N=6) and seronegative (N=17) vaccine recipients. The soluble gB-specific, PC-specific, and TB40/E virion-bound IgG antibody titers (ED50: effective dose 50) were determined via ELISA for both seropositive (B, D, F) and seropositive (C, E, G) vaccinees, respectively, at the indicated time points including the 1-year follow-up after last vaccination. The dotted line indicates the limit of detection (ED50=30). IgG binding activities to Merlin gB-transfected cells were evaluated for seronegative (H) and seropositive (I) vaccinees at the indicated time points via flow cytometry-based assay. p values from Wilcoxon matched-pairs signed rank test are reported. *p<0.05, **p<0.01, ***p<0.001.

**Table 1:** Vaccine participants in the mRNA-1647 vaccine cohort and the timepoints evaluated for humoral responses in current report.

| Participant ID | Age range (years) | Gender | Serostatus at baseline | Timepoints analyzed for humoral immune responses |  |  |  |
| --- | --- | --- | --- | --- | --- | --- | --- |
|  |  |  |  | Month 0 | Month 7 | Month 12 | Month18 |
| SP-01 | 16-20 | F<sup>a</sup> | S(+)<sup>c</sup><br>(n=6) | Y<sup>e</sup> | Y | Y | Y |
| SP-02 | 46-50 | M<sup>b</sup> |  | Y | Y | Y | Y |
| SP-03 | 36-40 | M |  | Y | Y | Y | ND |
| SP-04 | 41-45 | M |  | Y | Y | Y | ND |
| SP-05 | 36-40 | F |  | Y | Y | Y | Y |
| SP-06 | 26-30 | F |  | Y | Y | Y | Y |
| SN-01 | 41-45 | M | S(-)<sup>d</sup><br>(n=17) | Y | Y | Y | ND |
| SN-02 | 26-30 | F |  | Y | Y | Y | ND |
| SN-03 | 21-25 | F |  | Y | Y | Y | Y |
| SN-04 | 36-40 | F |  | Y | Y | Y | Y |
| SN-05 | 26-30 | M |  | Y | Y | Y | Y |
| SN-06 | 41-45 | F |  | Y | Y | Y | Y |
| SN-07 | 16-20 | M |  | Y | Y | Y | ND |
| SN-08 | 21-25 | F |  | Y | Y | Y | ND |
| SN-09 | 31-35 | M |  | Y | Y | Y | ND |
| SN-10 | 21-25 | F |  | Y | Y | Y | ND |
| SN-11 | 26-30 | F |  | Y | Y | Y | ND |
| SN-12 | 26-30 | F |  | Y | Y | ND<sup>f</sup> | ND |
| SN-13 | 46-50 | F |  | Y | Y | Y | ND |
| SN-14 | 46-50 | F |  | Y | Y | Y | ND |
| SN-15 | 36-40 | F |  | Y | Y | Y | ND |
| SN-16 | 41-45 | F |  | Y | Y | ND | Y |
| SN-17 | 26-30 | F |  | Y | Y | Y | Y |

F<sup>a</sup>: female; M<sup>b</sup>: male;
S(+)<sup>c</sup>: HCMV seropositive at baseline; S(-)<sup>d</sup>: HCMV seronegative at baseline
Y<sup>e</sup> :Yes, denotes the sample has been analyzed at this particular timepoint; ND<sup>f</sup>: not done due to unavailability of samples;

### IgG Binding Assays

Sera IgG binding to HCMV TB40/E virions, Towne gB, Towne gB AD2S1 peptide and VR1814 PC was measured by ELISA as previously described (9, 16, 21). Sera IgG binding to gB expressed on the cell surface was measured by flow cytometry as previously described (9, 16, 21). IgG and subclasses IgG1, IgG2, IgG3 binding to HCMV antigens (gB, gB AD-1, gB AD4, gB AD5, gB AD4+AD5, PC) and Fcγ receptors (FcγR1A, FcγR2A, FcγR2B, FcγR3A) and neonatal receptor (FcRN) were measured by binding antibody multiplex assay (BAMA) as described (9, 16, 21). Results were acquired on a Bio-Plex 200 system (Bio-Rad) and reported as mean fluorescence intensity (MFI).

### Functional Ab Assays

Neutralization was measured by employing HCMV immediate early-1 (IE-1) gene expression to quantify reduction in virus infection in fibroblast HFF-1 or epithelial ARPE-19 cells (9, 16, 21). ID_50_s were calculated using non-linear regression analysis (Sigmoidal, 4PL) using GraphPad Prism. ADCC was measured using cell-surface expression of CD107a as a marker for NK cell degranulation, as described (16, 21). Primary human NK cells were incubated with AD169-derivative BadrUL131-Y4/GFP-infected MRC-5 cell monolayers (22). The frequencies of CD107a+ live NK cells were determined by flow cytometry. For ADCP assays, an optimized amount of AD169r virions (1×10^3^ PFU/well) was conjugated to AF647 NHS ester before incubation with diluted sera (1:100). Virus-antibody immune complexes were spinoculated and incubated with THP-1 cells and stained with Aqua Live/Dead dye, and the percentage of AF647+ cells was reported for each sample based on the live, singlet population.

## RESULTS

### mRNA-1647 vaccination elicited durable HCMV-specific IgG responses

HCMV gB-specific IgG responses were significantly increased above baseline at seven months post-vaccination (peak timepoint) in seronegative vaccinees and elevated in seropositive vaccinees (Figure 1B, 1C). Similarly, PC-specific peak IgG responses were also significantly increased at month 7 in seronegative vaccinees and elevated in seropositive vaccinees (Figure 1D,1E). Peak gB-specific IgG responses in seronegative vaccinees were comparable to responses induced by natural infection, whereas PC-specific IgG responses were 10-fold higher than those induced by natural infection (Figure S2A-S1B). Peak gB- and PC-specific (Figure S3A-3F) IgG1, IgG2, and IgG3 responses were elicited in seronegative vaccinees, while only IgG1 responses were elevated above baseline in seropositive vaccinees (Figure S3A, 3D). gB- and PC-specific IgG responses persisted through one year after the last vaccination in seronegative vaccinees (Figure 1B, D). While IgG responses declined between month 7 to 12, no significant change was observed in vaccine-elicited IgG levels between month 12 to 18 (Figure 1B-1E). Interestingly, the relative avidity index (RAI) for gB-specific IgG declined significantly between month 7 and 12 only in seronegative vaccinees (Figure S4A-S4B). In contrast, PC-specific IgG maintained an equivalent RAI at both month 12 and 18 relative to month 7 in seronegative and seropositive vaccinees (Figure S4C-S4D).

Significant increases in HCMV-binding IgG levels against TB40/E (gB1 genotype) (21) were observed in seronegative, but not seropositive individuals, after vaccination (Figure 1F, 1G), though this was significantly lower than those elicited by natural infection (Figure S2C). Virion-binding IgG responses in seronegative vaccinees declined in magnitude (Figure 1F), but not in avidity (Figure S4E), between months 7 and 12 and returned to baseline by month 18. In seropositive vaccinees, there was no change in magnitude or avidity of virion-binding IgG after vaccination (Figure 1G, Figure S4F),

IgG binding to gB-transfected cells was previously identified as a correlate of protection against HCMV acquisition in the gB/MF59 vaccine trials (9). Thus, we measured IgG binding to mRNA-1647 vaccine strain-matched Merlin and heterologous gB1 genotype (Towne) gB-transfected cells for both vaccine populations. IgG responses to cell-associated Merlin and Towne gB were elicited in seronegative vaccinees (Figure 1H; Figure S5), and Merlin gB-transfected IgG binding responses remained high one year after the last vaccination. Only Towne cell-associated gB responses were significantly increased by vaccination in seropositive vaccinees (Figure 1I; Figure S5), demonstrating that mRNA-1647 was able to elicit broad IgG responses and boost pre-existing responses against the cell-associated conformation of gB.

### mRNA-1647 induced broad and long-lasting NAb responses

We measured NAb responses against homologous (Towne, gB1) and heterologous (AD169r, gB2) gB genotype strains in fibroblasts and epithelial cells. Low level fibroblast NAb responses against Towne (Figure 2A, 2B) and AD169r (Figure 2C, 2D) were elicited by month 7 after vaccination in seronegative mRNA-1647 vaccinees and were increased above baseline in seropositive vaccinees. In contrast, mRNA-1647 elicited potent epithelial NAb responses against AD169r in seronegative vaccinees at month 7 after vaccination, and >400-fold above the fibroblast neutralization titer (median ID50 40660 vs. 99). These responses were durable through one year after the last vaccination (Figure 2E). The magnitude of vaccine-elicited epithelial, but not fibroblast, NAb responses was significantly higher than those induced by natural infection (Figure S6A-S6C). This response was also increased by 55-fold compared to pre-existing responses in seropositive vaccinees and maintained through one year after the final vaccination (Figure 2F).

**Figure 2.**
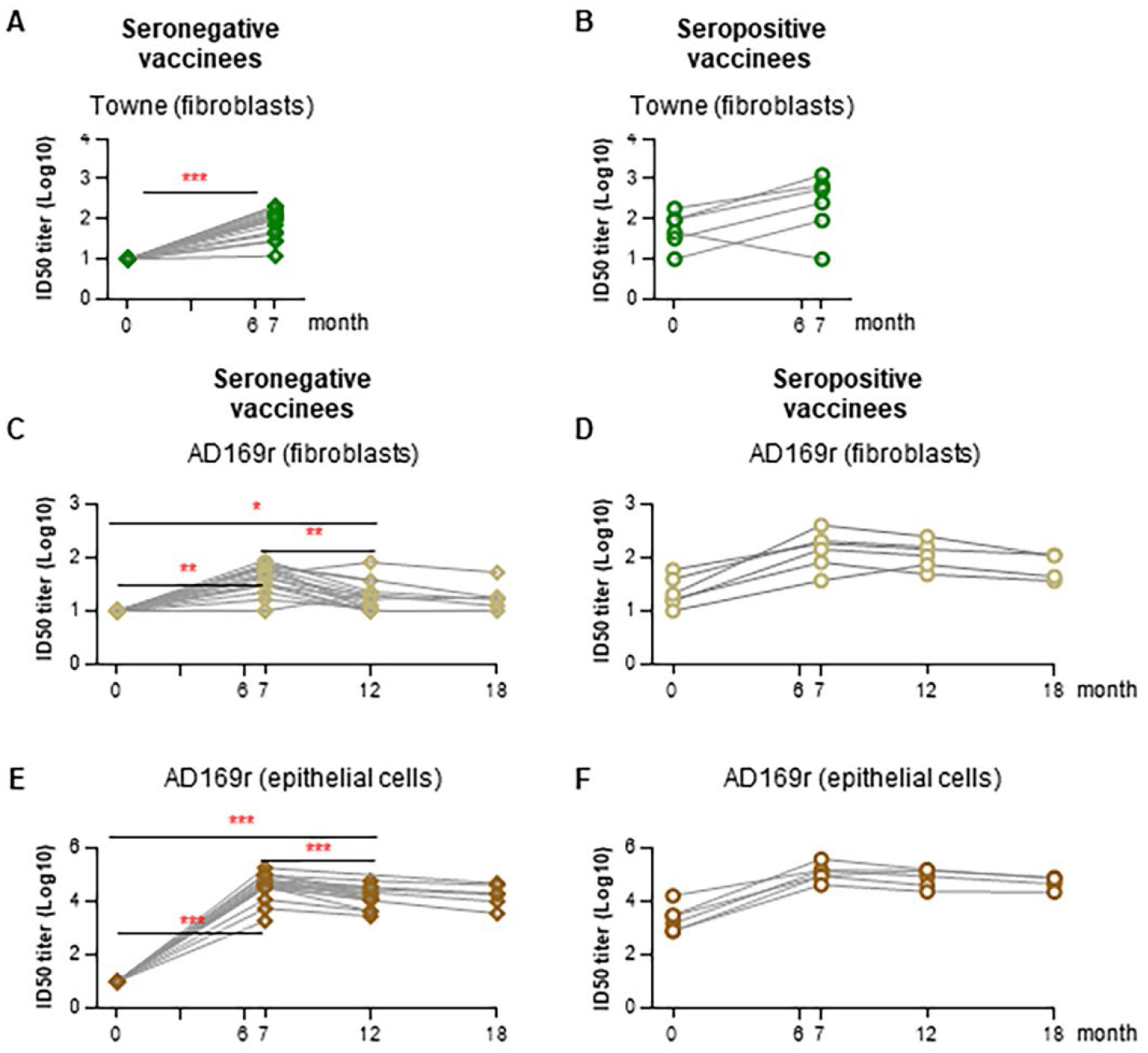
Durable neutralizing antibody responses elicited by the mRNA-1647 vaccine in both seropositive and seronegative vaccine recipients. Fibroblast neutralization reactivities against Towne virus in HFF-1 cells were compared before (month 0) and after (month7) vaccination for both seronegative (A) and seropositive (B) vaccinees. Similarly, fibroblast neutralization responses against AD169r virus in HFF-1 cells and epithelial neutralization antibody responses against AD169 in ARPE-19 cells were monitored before vaccination (month 0), month 7, 12 to 18 after vaccination for both seronegative (C, E) and seropositive (D, F) vaccinees, respectively, to assess durability. p values from Wilcoxon matched pairs signed rank test are reported. *p<0.05, **p<0.01, ***p<0.001.

IgG responses against gB antigenic domains (AD) were detectable against AD4, and AD4&AD5 (Figure S7B, S7D) in seronegative vaccinees at month 7 after mRNA-1647 vaccination, revealing an AD4 dominant IgG response that did not correlate with fibroblast NAb responses. In seropositive vaccinees, anti-AD4, AD4&D5, and AD2S1 responses were boosted by vaccination (Figure S7B, S7D-S7E), and these responses also did not correlate with fibroblast NAb responses. However, nAb responses against AD169r correlated strongly with AD4, AD4&AD5-specific IgG binding when seronegative and seropositive vaccinee responses were combined (Table S1).

### mRNA-1647 elicited Fc-mediated functional antibody responses

We next evaluated vaccine-induced antigen-specific Fc-mediated IgG responses. In all vaccinees, we found significant increases in both gB- and PC-specific (Figure 3A, 3B) IgG responses that bound to all tested FcRs (FcγR1A, FcγR2A, FcγR2B, FcγR3A and FcRN). Both ADCC and ADCP responses were significantly increased in seronegative vaccinees following vaccination but not in seropositive vaccinees (Figure 3C, 3D). ADCC and ADCP responses in seropositive vaccinees at baseline were significantly higher than peak responses for seronegative vaccines, suggesting that these responses are more potently elicited by infection than vaccination. AD169r virion-specific ADCC strongly correlated with all tested gB-specific, but not PC-specific, FcR-binding IgG levels in seronegative vaccinees (Table S2; Figure 3E-3F). When our analysis was expanded to the total vaccinated population, we observed strong correlation between ADCC and gB-specific FcR-binding IgG levels; particularly, gB-specific FcγR3A-binding (Figure 3F, r = 0.87, p <0.001), which is expressed on NK cells that mediate ADCC (23). Similarly, we observed positive correlation between AD169r virion-specific ADCP response and all evaluated gB- and PC-specific IgG FcR binding responses levels in seronegative vaccinees, and the correlation became stronger when the analysis was extended to the total vaccinated population (Table S2; Figure 3G-3H), in particular for gB-specific FcγR1A (r = 0.93, p <0.001) and FcγR2A (r = 0.93, p <0.001) (Figure 3H).

**Figure 3.**
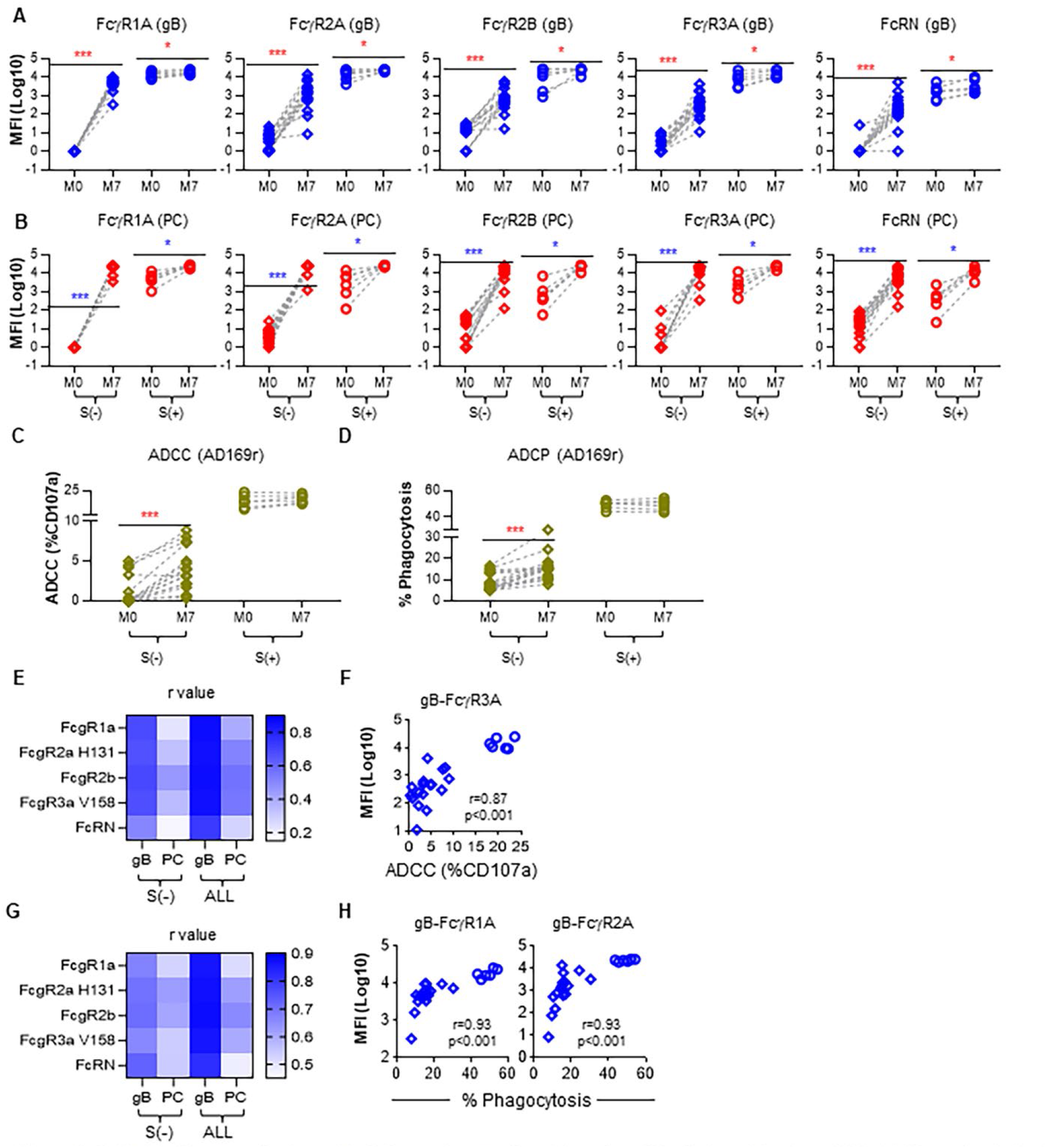
FcRs binding magnitude and IgG Fc-portion mediated functionalities induced by the mRNA-1647 vaccine in both seropositive and seronegative vaccinees. gB-specific (A) and PC-specific (B) IgG Fc antibody (FcgR1A, FcgR2A (H131), FcgR2B, FcgR3A (V158), and FcRN) responses were plotted for both seronegative and seropositive vaccinees before vaccination (month 0) and at the peak time point (month 7). Vaccine-induced IgG antibody Fc-mediated AD169r virion ADCC (C) and ADCP (D) responses were assessed for both seronegative and seropositive vaccinees. p values from Wilcoxon matched-pairs signed rank test are reported. Spearman correlation analysis between AD169r virion ADCC (E) and ADCP (G) responses and gB-specific and PC-specific IgG FcR binding magnitude with the spearman r value shown for seronegative only (S(−)) and all vaccinees (ALL) is reported. Correlation for ADCC and gB-specific FcR binding magnitude (F), ADCP and gB-specific (H) FcR binding magnitude for seronegative only (S(−)) and all vaccinees (ALL) is reported; S−: seronegative samples; ALL include seronegative (open diamond symbol) and seropositive samples (open circle symbol)). *p<0.05, **p<0.01, ***p<0.001.

### mRNA-1647 induced higher PC- than gB-specific IgG responses

To evaluate if balanced humoral responses were induced for both antigens encoded by the mRNA-1647 formulation, we compared vaccine-elicited PC- and gB-specific IgG responses. We found significantly higher PC- than gB-specific IgG responses in seronegative vaccines, including total IgG, IgG1, and IgG3 (Figure 4A-4D) responses. There was a trend towards higher PC- than gB-specific total IgG and IgG1 responses in seropositive vaccinees, but only PC-specific IgG3 responses were significantly higher (Figure 4E, 4F and 4H).

**Figure 4.**
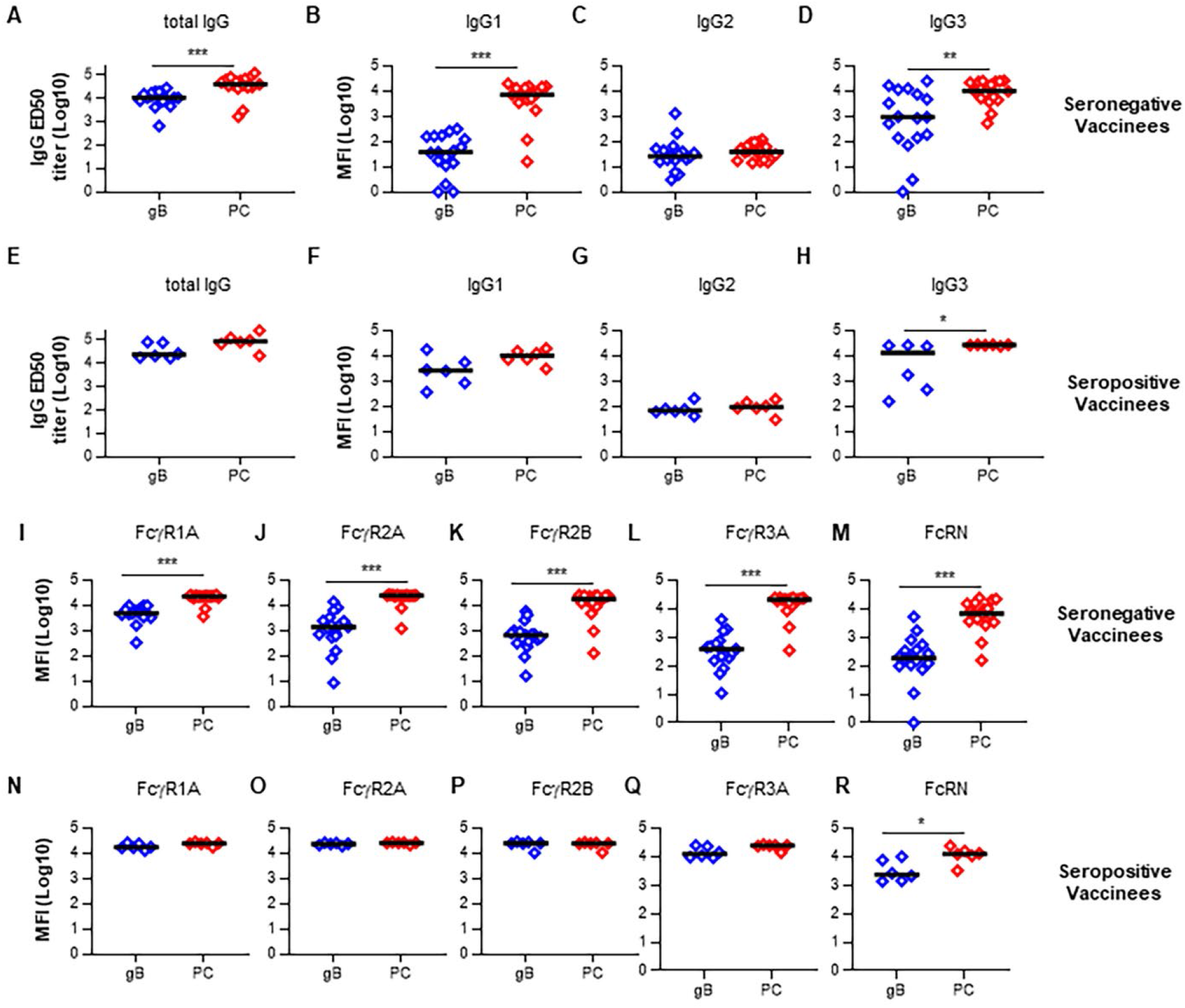
mRNA-1647 induced higher PC-specific IgG responses than gB-specific responses in the same formulation. The magnitude of mRNA-1647 vaccine-induced total IgG responses and IgG subclass (IgG1, IgG2 and IgG3) responses were compared between gB-specific and PC-specific responses for both seronegative (A-D) and seropositive (E-H) vaccinees at month 7. The total IgG responses were measured by ELISA, while the IgG subclass (IgG1, IgG2, and IgG3) responses were detected via BAMA. p values from Wilcoxon matched pairs signed rank test are reported. The level of IgG Fc antibody responses (FcγR1A, FcγR2A, FcγR2B, FcγR3A, and FcRN) were compared among gB and PC antigen components in the same vaccine formulation in both seronegative (I-M) and seropositive (N-R) vaccinees in mRNA-1647 vaccine cohort. p values from non-parametric t-tests are reported (Mann-Whitney). *p<0.05, **p<0.01, ***p<0.001.

PC-specific FcR-binding IgG responses were also higher in seronegative vaccinees compared to gB-specific responses (Figure 4I-4M); which was also observed when data was normalized to gB- and PC-specific IgG levels, (Figure S8) with the exception of FcγR1A binding. Further, PC-specific FcγR3A-binding IgG responses also trended higher than gB-specific responses in seropositive vaccinees (Figure 4Q). This difference may be underestimated as the PC-specific, but not gB-specific, responses were at the maximum limit of detection (Figure 4Q). Although no significant difference was found for FcγR1A, 2A and 2B IgG binding between PC-specific and gB-specific IgG responses in seropositive vaccinees, these responses were also at the maximum limit of detection (Figure 4N-4Q). FcRN-binding IgG responses that were PC-specific were elicited by month 7 in seropositive vaccinees (Figure 4R).

### mRNA-1647 elicited broad and potent NAb and higher ADCC responses, but lower gB-specific IgG and ADCP responses compared to gB/MF59 in seronegative vaccinees

We compared peak gB-specific IgG responses in HCMV-seronegative vaccinees at month 7 post-vaccination among two vaccine cohorts. The vaccines were delivered intramuscularly on a similar schedule, and included a gB1 genotype antigen (Figure 5A). gB/MF59 vaccine induced significantly higher gB-specific IgG responses compared to the mRNA-1647 vaccine, yet with similar avidity (Figure 5B, 5C). Vaccine-induced gB-specific IgG subclass responses, including IgG1 and IgG3 (Figure 5D-5F), were also significantly elevated in the gB/MF59 vaccine cohort, in which total IgG and IgG1 was higher than that induced by natural infection (Figure 5B, 5D). Interestingly, ratios of gB-specific IgG2/IgG1 and IgG3/IgG1 were higher in the mRNA-1647 vs gB/MF59 vaccine cohort (Figure 5E-5F) suggesting distinct vaccine-induced IgG subclass patterns. Despite the inclusion of 2 major glycoprotein complexes in mRNA-1647, gB/MF59 vaccination elicited higher virion-binding IgG against TB40/E in terms of both magnitude (Figure 5G) and avidity (Figure 5H).

**Figure 5.**
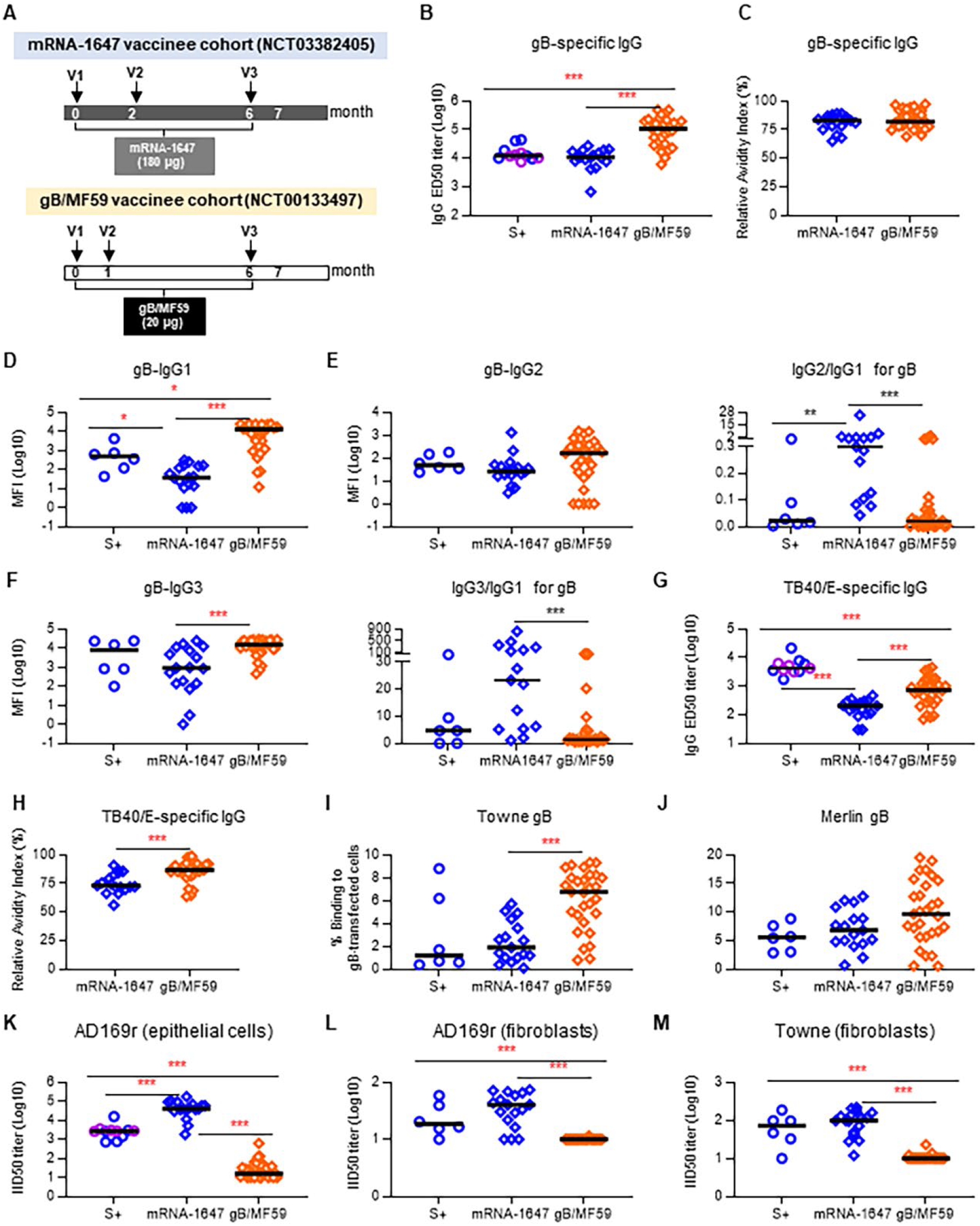
Comparison of gB-specific peak IgG binding antibody and neutralizing antibody responses. (A): Cartoons illustrating the immunization schedules for mRNA-1647 and gB/MF59 vaccine cohorts for vaccine-induced gB-specific antibody peak responses comparison at month 7. Soluble gB-specific total IgG antibody titers (B) relative avidity index (RAI) (C) and IgG subclass IgG1 (D), IgG2 (E), and IgG3 (F) responses were compared between the two vaccine cohorts. TB40/E virion-bound IgG titers (G) and RAI (H) were also compared between the two vaccine cohorts. Similarly, Towne gB (I) and Merlin gB (J) cell-associated IgG antibody responses were compared among the two vaccine cohorts. The epithelial neutralizing antibody against AD169r (K) in ARPE-19 cells and fibroblast neutralizing antibody against AD169r (L) and against Towne (M) in HFF-1 cells were compared between mRNA-1647 and gB/MF59 vaccine cohorts. Additionally, the HCMV infected individuals were also included for the comparison. Open blue circle denotes the seropositive samples at baseline from the mRNA-1647 vaccine cohort while open purple circle denotes the in-house HCMV seropositive subjects in some assays to increase the statistical power. But the statistical analysis indicated the increased sample # didn’t change the conclusion. P values from non-parametric t-tests are reported (Mann-Whitney). *p<0.05, **p<0.01, ***p<0.001.

gB/MF59 immunization also elicited higher cell-associated autologous Towne gB-specific IgG responses than the mRNA-1647 vaccine (Figure 5I). IgG against cell-associated Merlin gB (Figure 5J) that trended higher in the gB/MF59 cohort than that induced by mRNA-1647. Finally, no difference in gB AD-specific IgG responses was found among gB/MF59 and mRNA-1647 vaccine cohorts (Figure S9A-S9E, Table 2).

**Table 2:** Comparison of humoral responses in the mRNA-1647 vaccine cohort and gB/MF59 vaccine cohort.

| Humoral Response | mRNA-1647<br>Seronegative<br>vaccinees (n = 17) | gB/MF59<br>Seronegative vaccinees<br>(n = 29) | FDR-adjusted<br>p-value |
| --- | --- | --- | --- |
|  | Median (Range) | Median (Range) |  |
| **IgG binding** |  |  |  |
| *Soluble gB* |  |  |  |
| gB-specific (Log<sub>10</sub>ED<sub>50</sub>) | 4.03 (2.82, 4.44) | **5.03 (3.78, 5.69)** | < 0.001 |
| Avidity (%RAI) | 83.00 (65.00, 89.00) | 81.91 (68.99, 97.27) | 0.402 |
| *Virion-specific* |  |  |  |
| TB40e-specific (Log<sub>10</sub>ED<sub>50</sub>) | 2.31 (1.48, 2.67) | **2.86 (1.85, 3.66)** | < 0.001 |
| Avidity (%RAI) | 73.10 (56.10, 90.80) | **86.72 (63.58, 98.87)** | 0.001 |
| *Cell-associated gB (%binding)* |  |  |  |
| Towne | 1.90 (0.10, 5.70) | **6.76 (0.79, 9.33)** | < 0.001 |
| Merlin | 6.81 (0.71, 12.75) | 9.64 (0.54, 19.50) | 0.116 |
| **gB-specific IgG subclass (Log<sub>10</sub>MFI)** |  |  |  |
| IgG1 | 1.57 (0.00, 2.50) | **4.10 (1.09, 4.39)** | < 0.001 |
| IgG2 | 1.41 (0.48, 3.11) | 2.22 (0.00, 3.17) | 0.094 |
| IgG3 | 2.96 (0.00, 4.39) | **4.18 (2.65, 4.46)** | < 0.001 |
| **Neutralization (Log<sub>10</sub>ID<sub>50</sub>)** |  |  |  |
| Towne (fibroblast) | **2.00 (1.08, 2.34)** | 1.00 (1.00, 1.37) | < 0.001 |
| AD169r (fibroblast) | **1.60 (1.00, 1.87)** | 1.00 (1.00, 1.06) | < 0.001 |
| AD169r (epithelial) | **4.61 (3.26, 5.24)** | 1.20 (1.00, 2.79) | < 0.001 |
| **gB AD mapping (Log<sub>10</sub>MFI)** |  |  |  |
| AD-1 | 1.1 (0.0, 1.7) | 1.0 (0.0, 2.4) | 0.892 |
| AD-2 site 1 (AUC) | -0.6 (-0.8, 0.5) | -0.5 (-0.7, 0.4) | 0.3 |
| AD-4 | 1.9 (0.0, 3.0) | 1.5 (0.0, 2.5) | 0.686 |
| AD-5 | 0.0 (0.0, 1.9) | 1.0 (0.0, 2.3) | 0.3 |
| AD-4&5 | 1.6 (0.0, 2.9) | 1.7 (0.0, 2.8) | 0.686 |
| **gB-specific FcR binding (Log<sub>10</sub>MFI)** |  |  |  |
| FcR<sub>γ</sub>R1A | 3.68 (2.51, 3.98) | 3.72 (2.50, 4.10) | 0.826 |
| FcR<sub>γ</sub>R2A | 3.14 (0.93, 4.14) | **4.36 (2.26, 4.40)** | < 0.001 |
| FcR<sub>γ</sub>R2B | 2.81 (1.20, 3.76) | **4.35 (2.10, 4.41)** | < 0.001 |
| FcR<sub>γ</sub>R3A | 2.58 (1.04, 3.62) | **4.27 (2.64, 4.40)** | < 0.001 |
| FcRN | 2.26 (1.00, 3.72) | **3.72 (2.11, 4.39)** | < 0.001 |
| **Fc-mediated effector functions** |  |  |  |
| ADCC (% CD107A) | **3.20 (0.30, 8.90)** | 1.60 (0.00, 16.80) | 0.032 |
| ADCP (% Phagocytosis) | 15.20 (7.80, 30.50) | **20.20 (13.10, 47.10)** | < 0.001 |

p<0.05 (bolded) is considered significant. Underlined values represent significantly higher responses.
FDR: False discovery rate, RAI: Relative Avidity Index, MFI: Mean Fluorescent Intensity.

Despite the lower gB-specific IgG responses compared to those elicited by gB/MF59, mRNA-1647 vaccine induced higher heterologous virus (AD169r) NAb responses in both epithelial (Figure 5K) and fibroblast (Figure 5L) cells, including fibroblast NAb responses against the gB/MF59 homologous Towne virus (Figure 5M), indicating that mRNA-1647 vaccine-induced PC-specific antibody responses may contribute to the fibroblast-tropic NAb responses (24). Additionally, the gB/MF59 vaccine induced higher gB-specific IgG FcγR2A, 2B, 3A and FcRN binding responses compared to that of mRNA-1647, yet similar levels of binding to FcγR1A (Figure 6A–6E). When normalized to gB-specific IgG responses, the gB/MF59 vaccinees still had higher FcγR2B, 3A, and FcRN, but lower FcγR1A (Figure S10C-S10E and S10A), binding responses (Figure S10B). Finally, mRNA-1647 vaccine induced higher ADCC, yet lower ADCP (Figure 6F, 6G) responses compared to the gB/MF59 vaccine. No significant correlation was found between tested FcR binding and ADCC/ADCP responses in the gB/MF59 cohort (Table S3). The humoral response comparison between the vaccine cohorts is summarized in Table 2.

**Figure 6.**
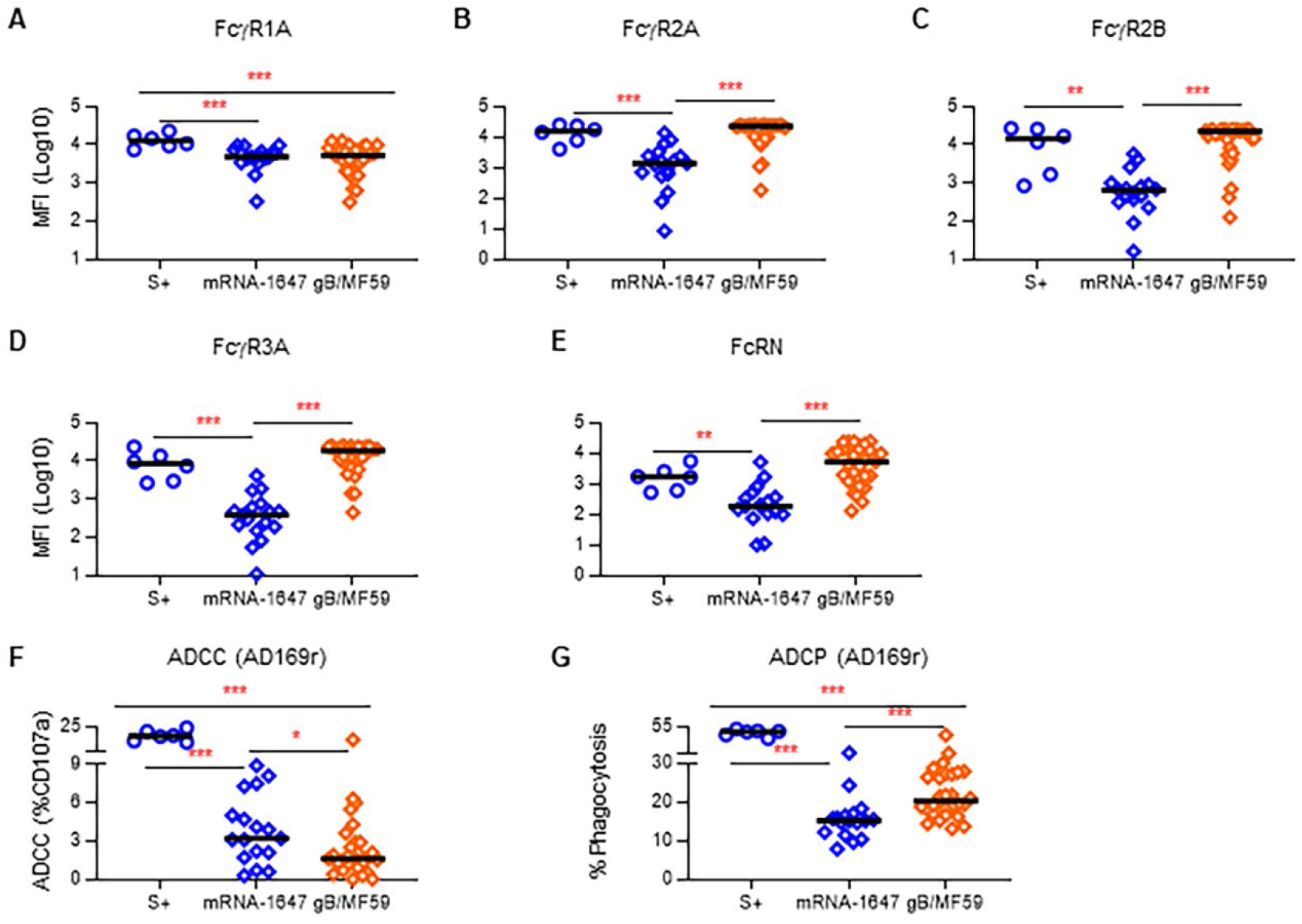
gB/MF59 vaccine elicited higher peak FcRs and ADCP but lower ADCC functional antibody responses than the mRNA-1647 vaccine candidate. gB-specific FcgR1A (A), 2A (B), 2B (C), 3A (D), and FcRN (E) antibody responses were compared between the two vaccine cohorts indicative of overall higher-level responses except for FcgR1A response in the gB/MF59 vaccine cohort; AD169r virion ADCC (F) and ADCP (G) effector antibody responses were compared between two vaccine cohorts suggesting elevated ADCC responses, but lower ADCP responses were found in mRNA-1647 cohort. Additionally, the HCMV infected individuals were also included in the comparison. Open blue circle denotes the seropositive samples at baseline from the mRNA-1647 vaccine. p values from non-parametric t-tests are reported (Mann-Whitney). *p<0.05, **p<0.01, ***p<0.001.

## DISCUSSION

In this report, we demonstrate that mRNA-1647 HCMV vaccine induced potent and durable HCMV-specific IgG responses in seronegative participants in a first-in-human vaccine trial, mirroring preclinical immunogenicity results (19, 20). It also boosted pre-existing HCMV-specific IgG responses in seropositive vaccinees. mRNA-1647 elicited broad and potent neutralization as well as Fc-mediated effector functions, including robust ADCC responses. These features support its advancement into ongoing late-stage clinical trial assessing its efficacy for prevention of HCMV acquisition in seronegative women.

Humoral immune correlates of protection against HCMV acquisition or vertical transmission are still being investigated (8); hence broad and polyfunctional humoral immunity is desirable. Interestingly, gB-specific IgG responses in mRNA-1647 vaccinees was considerably lower than that of the PC-specific IgG response and that elicited by the gB/MF59 vaccine, indicating potential room for improvement of the immunogenicity of the gB antigen. The lower magnitude of gB vs PC-specific IgG responses could be in part be due to the lower total amount of gB vs PC mRNA in the vaccine formulation (gB vs. PC: 30μg vs.150μg), suggesting re-formulation of the vaccine could lead to more balanced responses. Alternative approaches could combine mRNA and protein antigens in a co-delivery strategy, similar to that applied other vaccine platforms (28, 29). Of note, PC was recently shown to be dispensable for cCMV transmission in NHPs (25); yet, the impact of the addition of PC to an HCMV vaccine on efficacy is yet unclear, but will be informed by the ongoing mRNA-1647 trial.

While neutralization responses were absent or minimal in gB/MF59 vaccinees, the PC-specific IgG and neutralization responses elicited by mRNA-1647 were quite robust (Table 2). Yet, the magnitude of gB-specific IgG responses elicited by mRNA-1647, assayed against the heterologous Towne gB protein, were significantly lower than that elicited by the gB/MF59 vaccine. The elicited IgG binding to cell-associated Towne gB, the immune correlate of protection in the gB/MF59 vaccine trials (9), was also lower in the mRNA-1647 group. However, IgG binding to cell-associated Merlin gB, which is strain matched to the mRNA-1647 vaccine, was equivalent among the vaccinees (Figure 5J). Further, ADCC responses elicited by mRNA-1647 were more robust than that of the gB/MF59 vaccine, while ADCP responses were lower. Both effector responses have recently been associated with reduced cCMV transmission risk (16, 17). The lower ADCP responses elicited by mRNA-1647 vs gB/MF59 vaccine could be attributable to the distinct gB-specific FcR binding (Figure S10) or IgG subclass ratios (Figure 4D-F), but also indicate that PC-specific responses may contribute little to this response. Conversely, the higher ADCC response in mRNA-1647 vs gB/MF59 vaccine indicates that the PC is an important target of ADCC.

The expected differences in gB conformation can potentially explain differences in the vaccine-elicited functional responses; gB/MF59 vaccine has a truncated transmembrane domain and a mutated furin cleavage site (9), while the mRNA-1647 encodes an intact gB protein (19, 20) that may be expressed in the context of a cell membrane. Yet, the gB AD-specific IgG responses were not distinct. One report demonstrated that a Towne ectodomain gB antigen induced better immunogenicity than the full length gB antigen when delivered in the same viral vector (26). Distinct vaccinee populations may also be a confounding factor for comparison between the cohorts, as previously described in immunity elicited by gB/MF59 (9) and natural infection (27). The gB/MF59 vaccinees were a lower age range (12-17) than that of mRNA-1647 (18-49) and only female gender, though most vaccinees in the mRNA-1647 cohort were female (76%).

Durable IgG responses elicited by mRNA-based HCMV vaccine mirrors results of the preclinical models including mice, NHPs (20), and rabbits (30). Yet, it is critical to assess durability at longer intervals given the mRNA-167 vaccine strategy to vaccinate in adolescence for protection against HCMV through reproductive years. Our observation of humoral immune responses lasting over 1 year after mRNA-1647 vaccination may stem from long-lived germinal center reactions upon mRNA-LNP vaccination as observed in SARS-CoV2 mRNA-LNP-immunized mice (31) and humans (32). Yet, due to sample availability, we were unable to compare response durability with that of gB/MF59 vaccine, which is a limitation of this study.

The immune responses required to prevent cCMV transmission may be distinct from those to prevent acquisition. While prevention of mucosal HCMV acquisition may be a challenge, rapid control of viral replication could be effective in eliminating transmission through vaccine-induced Fc-mediated antibody functions such as ADCC (33), ADCP (34) or T cell responses (3). Though T cell responses were not evaluated here, prior studies demonstrated that the mRNA-1647 vaccine is able to elicit potent cell-mediated immunity (20).

The advantages of mRNA-LNP vaccine, such as rapid development and flexible re-formulation provide an opportunity to iteratively redesign vaccine immunogens. The impact of the addition of PC and lower magnitude gB-specific IgG responses between the mRNA-1647 and the gB/MF59 vaccine on HCMV acquisition will be informed by the ongoing efficacy trial (NCT05085366), an important benchmark for the HCMV field and the immunogenicity requirements of a successful HCMV vaccine.

## Data Availability

All data produced in the present work are contained in the manuscript

## Author Contributions

**Investigators** were X.H., K.P.K., S.H., S.M.V., J.A.J, H.W., I.G.M., M.C., and J.P., E.B.W., K.M.E., D.I.B., and S.R.P. **Study design** was carried out by X.H., and S.R.P. **The trial was managed** by J.H., M.K., K. M. E., D.I.B., K.W., A.C. **Data analysis and interpretation** was carried out by X.H., K.P.K., S.H., S.M.V., J.A.J, H.W., J.P., C.A., L.M.G. and S.R.P. **Manuscript** was drafted by X.H. and edited by K.P.K. and S.R.P. All authors had full access to the presented data, provided critical input during manuscript preparation, and approved the manuscript for submission. We also thank Ms. Carolyn Weinbaum and other members of Dr. Sallie Permar’s lab for their support.

