## Supplementary Material for "Human Cytomegalovirus mRNA-1647 Vaccine Candidate Elicits Potent and Broad Neutralization and Higher Antibody-Dependent Cellular Cytotoxicity Responses than that of the Partially Effective gB/MF59 Vaccine"

**Supplementary Methods**

***Study Design and Participants***

We assessed humoral responses in 23 healthy adult participants from a phase I clinical trial (December 2017 – October 2020) immunized with 3 doses of mRNA-1647 (180 μg) vaccine at month 0, 2 and 6 (mRNA-1647 cohort; NCT03382405). Sera samples were collected pre-immunization (month 0) and one month post dose 3, including 6 previously exposed to HCMV (seropositive) and 17 naïve to HCMV infection (seronegative) samples at baseline. To evaluate durability of humoral responses, we obtained sera samples 12- and 18-months post recruitment for a partial group of donors (Figure 1A and Table 1). Samples from mRNA-1647 cohort were provided by Moderna.

To compare vaccine-induced HCMV gB-specific humoral responses, we also evaluated immune responses in 29 HCMV seronegative female adolescent donors from an MF59 adjuvanted subunit gB vaccine clinical trial (gB/MF59 cohort; NCT00133497) (9). Participants were immunized with 3 doses of gB/MF59 vaccine (20 μg) at month 0, 1 and 6 (Figure 5A), and we obtained samples one month post dose 3. Sera samples from gB/MF59 cohort were previously obtained from phase II vaccine clinical trial (Sanofi/Novartis). Institutional review board (IRB) approval was obtained for the gB/MF59 cohort from the Cincinnati Children’s Hospital Medical Center, and samples were provided by the NIAID Vaccine Treatment and Evaluation Unit (grant #5R21 AI136556). For the mRNA-1647 study conducted by Moderna, the study protocol, amendments, and informed consent form were reviewed and approved by the Advarra IRB. Written informed consent was obtained from all participants prior to study recruitment and procedures. The study protocol adhered to all applicable national, state, and local laws or regulations, the principles of the International Council for Harmonisation harmonized tripartite guideline E6(R2): Good Clinical Practice and of the Declaration of Helsinki. All mRNA-1647 samples transferred to Duke and WCM from Moderna were de-identified, and the analysis of de-identified samples was exempt from human subject’s research designation by the IRB’s of Duke and WCM. The information about the study participants is summarized in Table 1. The gB/MF59 vaccinees being a lower age range (13 to 17, median 15) than that of mRNA-1647 (18-49, median 30 for seronegative and median 38 for seropositive participants) (Figure S1).

A report regarding the safety, reactogenicity and cellular immune responses for mRNA-1647 vaccine is being submitted for publication.

***Study Products*** ***and*** ***Study Procedure***

mRNA-1647 is formulated in a single dose comprised of equal mass amount (30μg) of 5 mRNA molecules (UL75, UL115, UL128, UL130, UL131A) to encode gH, gL, UL128, UL130, UL131A subunits (Figure S11) to form the intact PC complex and an additional mRNA molecule (UL55, 30μg) to express the full-length (FL) envelope glycoprotein B (Figure S12), encapsulated with a proprietary ionizable cationic LNP from Moderna (19, 20). All six mRNA molecules were chemically N1-methyl-pseudouridine-modified (18, 35, 36) and both PC and gB were from HCMV reference strain Merlin with gB genotype 1 (gB1) (19-21).

For the gB/MF59 vaccine, 20 μg of soluble gB protein was formulated with oil-in-water emulsion adjuvant-MF59. gB antigen from HCMV Towne strain with gB1 genotype (Figure S12) (21) was transmembrane truncated and furin cleavage site mutated to enhance antigen expression and secretion (9).

***Antigen-specific IgG Binding Antibody and Relative Avidity Measurement by Enzyme Linked Immunosorbent Assays (ELISA)***

Sera IgG binding activity to HCMV TB40/E whole virions, FL Towne gB (gift from Sanofi Pasteur), Towne gB AD2S1 peptide and VR1814 PC were measured by ELISA as previously described (9, 16, 21). Briefly, 384-well plates were coated with virions at an optimized concentration of plaque-forming units (PFU) per well (100 PFU/well for TB40/E) or with 2 μg/ml of antigen protein/peptide per well. Data was acquired via Bioteck SL and reported as median effective dose 50 (ED50) titers or area under curve (AUC) (for AD2S1). Avidity was reported as Relative Avidity Index (RAI), calculated as the ratio of the OD450 of wells treated for 5 minutes with 7M urea to that of paired 1xPBS-treated wells.

***gB-transfected Cell IgG Binding Measured by Flow Cytometry***

Sera IgG binding to gB expressed on the cell surface was measured as previously described (9, 16, 21). Briefly, Human embryonic kidney HEK 293T/17 cells were co-transfected with DNA plasmids expressing GFP and full-length gB open reading frame (HCMV Towne and Merlin) for 48 hours and then incubated at 37ºC with 1:6250 diluted sera samples. Cells were stained with Live/Dead Fixable Near-IR Dead Cell Stain, followed by PE-conjugated goat-anti-human IgG Fc staining, and fixed with 10% formalin prior to acquisition via high throughput sampler (HTS) on the flow cytometer (Fortessa; BD). The frequency of PE+ cells was reported for each sample based on the live, singlet, GFP+ population.

***Antigen-specific IgG-binding Measurement by Binding Antibody Multiplex Assay (BAMA)***

HCMV-specific IgG and subclass IgG1, IgG2, IgG3 and Fc gamma receptors (FcγR1A, FcγR2A, FcγR2B, FcγR3A) and neonatal receptor (FcRN) binding to HCMV antigens (FL gB, gB AD-1, gB AD4, gB AD5, gB AD4 + AD5, PC) were measured by multiplex assay as previously described (9, 16, 21). Antigens were covalently coupled to fluorescent polystyrene beads (Luminex) and incubated with diluted sera samples (1:500 dilution for IgG1, 1:40 dilution for IgG2 and IgG3, 1:500 dilution for FcγR1A, FcγR2A, FcγR2B, FcγR3A, FcRN). Antibody binding was detected using phycoerythrin-conjugated goat-anti-human IgG secondary antibody (2 µg/mL, Southern Biotech). Results were acquired on a Bio-Plex 200 system (Bio-Rad) and reported as mean fluorescence intensity (MFI).

***Neutralization Assay***

Sera samples from vaccine participants or naturally infected individuals were heated at 56 °C for 30 min prior to assay. Neutralization was measured by employing HCMV immediate early-1 (IE-1) gene expression to quantify reductions in virus infection in HFF-1, MRC-5 or ARPE-19 cells for fibroblast and epithelial neutralization (9, 16, 21), respectively. Briefly, 6000 cells were seeded per well of a 384-well plate and incubated at 37 °C overnight. 7 μl of virus (MOI=1) was incubated with serial 3-fold dilutions of sera samples/IgG MAbs in a total volume of 60 μl for 1 hour at 37 °C in 384-well flat-bottom culture plates starting with 10 or 40 dilutions. Then, 25 μl of sera/virus mixture was added to cells in duplicate. One set of control wells received cells + virus (virus control), while another set received cells only (cell control). Following 24- or 48-hours incubation at 37 °C, 20 μl of 10% formalin was added to fix the cells for 10 mins at room temperature (RT), followed by staining HCMV IE-1 positive cells using an anti-IE-1 antibody for 1 hour. Next, the cells were stained with IgG-AF488 for 1 hour at RT, followed by nuclear staining with DAPI for 10 min at RT. Images were acquired using ImageXpress Pico Automated Cell Imaging System (Molecular Devices). Infected cells are calculated as a percentage of AF488 positive cells relative to total number of cells determined by DAPI staining. The 50% inhibitory dose (ID50) or concentration (IC50) was defined as the reciprocal of the sera dilution or concentration of IgG MAbs that caused a 50% reduction in infected cells compared to virus control (21). ID50s were calculated using non-linear regression analysis (Sigmoidal, 4PL) using GraphPad Prism. Raw image files are included for different conditions to show determination of ID50 or IC50 (Figure S13).

***HCMV Antibody-dependent cell-mediated Cytotoxicity Assay (ADCC)***

Cell-surface expression of CD107a was used as a marker for NK cell degranulation similar to previously described methods (16, 21). Primary human NK cells were added to wells containing AD169-derivative BadrUL131-Y4/GFP-infected MRC-5 cell monolayers (22). Diluted sera samples were added with Brefeldin A (GolgiPlug, 1 μl/mL, BD), monensin (GolgiStop, 4μl/6mL, BD), and anti-CD107a-FITC (BD, clone H4A3). After a 6-hour incubation, NK cells were washed and stained with a viability dye, anti-CD56-PE/Cy7 (BD, clone NCAM16.2), and anti-CD16-PacBlue (BD, clone 3G8). The frequencies of CD107a+ live NK cells were determined by flow cytometry. Final data representing specific activity were determined by subtraction of non-specific activity observed in assays performed with mock-infected cells.

***HCMV Virion Antibody-dependent cell-mediated Phagocytosis Assay (ADCP)***

Optimized amount of AD169r virions (1x10^3^ PFU/well) was conjugated to AF647 NHS ester prior to incubation with diluted sera samples (1:100). Then, virus-antibody immune complexes were centrifuged with 50,000 THP-1 cells for 1 hour at 1200 ×g and incubated at 37 °C for one hour. Cells were stained with Aqua Live/Dead stain, fixed with 10% formalin, and washed prior to acquisition on the flow cytometer (Fortessa; BD) using the HTS as previously described (16, 21). The percentage of AF647+ cells was reported for each sample based on the live, singlet population.

***Statistical Analysis****:*

Statistical analysis of immunological data and basic graphical delineation were performed using GraphPad Prism version 9.4.0 (GraphPad Software, Inc, La Jolla, CA) and R (4.2.2) based on a Spearman rank correlation test, a paired Wilcoxon test, or Mann-Whitney U test, where appropriate; *P* < 0.05 was considered significant. P-values are adjusted for multiple comparisons using the Benjamini and Hochberg (FDR) method which controls for the false discovery rate. In the FDR method, p values are ranked in an ascending array and multiplied by m/k where k is the position of a p value in the sorted vector and m is the number of independent tests. Neutralization titers (ID50) below the limit of detection (1:10 dilution) were assigned a value of 10 for statistical analysis purposes.

**Supplementary Table 1 summary of correlation analysis between neutralization titers and AD IgG-binding magnitude in mRNA-1647 cohort**

|  | Fi Neut-Towne | | | | Fi Neut-AD169 | | | | Epi Neut-AD169 | | | |
| --- | --- | --- | --- | --- | --- | --- | --- | --- | --- | --- | --- | --- |
|  | seronegative (n=17) | | total (n=23) | | seronegative (n=17) | | total (n=23) | | seronegative (n=17) | | total (n=23) | |
|  | **R** | **p** | **r** | **p** | **r** | **p** | **r** | **p** | r | p | r | p |
| gB AD-1 | -0.338 | 0.411 | 0.234 | 0.441 | -0.290 | 0.441 | 0.058 | 0.871 | -0.097 | 0.803 | 0.210 | 0.503 |
| gB AD5 | 0.096 | 0.803 | 0.489 | 0.072 | 0.244 | 0.503 | 0.437 | 0.151 | 0.452 | 0.207 | 0.563 | **0.036** |
| gB AD4 | 0.125 | 0.761 | 0.281 | 0.411 | 0.287 | 0.441 | 0.559 | **0.041** | 0.377 | 0.326 | 0.561 | **0.036** |
| gB AD4&AD5 | 0.137 | 0.745 | 0.339 | 0.293 | 0.246 | 0.503 | 0.595 | **0.036** | 0.579 | 0.068 | 0.683 | **<0.001** |
| AD2S1 | -0.025 | 0.926 | 0.159 | 0.602 | -0.028 | 0.926 | 0.265 | 0.441 | 0.059 | 0.871 | 0.248 | 0.441 |

**Supplementary Table 2 summary of correlation analysis between ADCC/ADCP responses and vaccine antigen-specific IgG FcR interaction in mRNA-1647 cohort**

|  | ADCC | | | | ADCP | | | | |
| --- | --- | --- | --- | --- | --- | --- | --- | --- | --- |
|  | seronegative (n=17) | | total (n=23) | | | seronegative (n=17) | | total (n=23) | |
| **gB** | r | adjusted p | r | adjusted p | | r | adjusted p | r | adjusted p |
| FcγR1a | 0.682 | **0.006** | 0.861 | **<0.001** | | 0.669 | **0.006** | 0.860 | **<0.001** |
| FcγR2a H131 | 0.661 | **0.007** | 0.851 | **<0.001** | | 0.699 | **0.005** | 0.871 | **<0.001** |
| FcγR2b | 0.686 | **0.005** | 0.859 | **<0.001** | | 0.706 | **0.005** | 0.876 | **<0.001** |
| FcγR3a V158 | 0.667 | **0.006** | 0.849 | **<0.001** | | 0.659 | **0.007** | 0.860 | **<0.001** |
| FcRN | 0.505 | **0.047** | 0.727 | **<0.001** | | 0.725 | **0.003** | 0.817 | **<0.001** |
| **PC** | r | adjusted p | r | adjusted p | | r | adjusted p | r | adjusted p |
| FcγR1a | 0.227 | 0.391 | 0.395 | 0.073 | | 0.527 | **0.038** | 0.512 | **0.019** |
| FcγR2a H131 | 0.331 | 0.210 | 0.514 | **0.018** | | 0.623 | **0.013** | 0.620 | **0.005** |
| FcγR2b | 0.452 | 0.078 | 0.564 | **0.009** | | 0.605 | **0.015** | 0.655 | **0.003** |
| FcγR3a V158 | 0.348 | 0.190 | 0.549 | **0.012** | | 0.539 | **0.034** | 0.599 | **0.006** |
| FcRN | 0.175 | 0.501 | 0.277 | 0.212 | | 0.542 | **0.033** | 0.480 | **0.028** |

**Supplementary Table 3 Summary of correlation analysis between ADCC/ADCP and vaccine antigen-spcific FcR interaction in the gB/MF59 cohort**

|  | ADCC | | ADCP | |
| --- | --- | --- | --- | --- |
|  | seronegative (n=29) | | | |
| **gB** | r | adjusted p | r | adjusted p |
| FcγR1a | -0.025 | 0.992 | 0.117 | 0.992 |
| FcγR2a H131 | 0.029 | 0.992 | 0.217 | 0.992 |
| FcγR2b | 0.016 | 0.992 | 0.189 | 0.992 |
| FcγR3a V158 | 0.002 | 0.992 | 0.178 | 0.992 |
| FcRN | -0.111 | 0.992 | 0.013 | 0.992 |

**Supplementary Figure Legends**

**Supplementary Figure 1. Age distribution among the vaccine cohort population.**
The age by year for vaccine recipients from both vaccine cohorts was plotted.

**Supplementary Figure 2. Comparison of HCMV-specific IgG antibody response among mRNA-1647 seronegative vaccinees and natural infected individuals.**
gB-specific (A), PC-specific (B) and TB40/E whole virion (C) IgG antibody responses in mRNA-1647 seronegative vaccinees at the month 7 and infected individuals were compared.

Open blue/red/green circle denotes the seropositive samples at baseline from the mRNA-1647 vaccine cohort while open purple circle denotes the in-house HCMV seropositive to increase the statistical power. But the statistical analysis indicated the increased sample # didn’t change the conclusion. P values from non-parametric t-tests are reported (Mann-Whitney). *p<0.05, **p<0.01, ***p<0.001.

**Supplementary Figure 3. mRNA-1647 induced antigen-specific IgG subclass responses in both HCMV seropositive and seronegative vaccinees.**
IgG subclass IgG1, IgG2, IgG3 antibodies were analyzed for both gB-specific (A, B, C) and PC-specific (D, E, F) responses, respectively, at month 0 and month 7 by BAMA. MFI for IgG subclasses was plotted for comparison for both seropositive and seronegative vaccinees at month 0 and month 7 post vaccination. p values from Wilcoxon matched-pairs signed rank test are reported. *p<0.05, **p<0.01, ***p<0.001.

**Supplementary Figure 4. Relative avidity of mRNA-1647 induced antigen-specific IgG responses.**

The relative avidity index of soluble gB-specific (A, B), soluble PC-specific (C, D), and TB/40E virion-bound (E, F) IgG antibodies was calculated for both seronegative (A, C, E) and seropositive (B, D, F) vaccinees, respectively, from day 0, month 7, 12, and 18 to evaluate the long-term vaccine-induced IgG avidity. p values from Wilcoxon matched-pairs signed rank test are reported. *p<0.05, **p<0.01, ***p<0.001.

**Supplementary Figure 5. Towne gB-transfected binding IgG responses induced by mRNA-1647 vaccine candidate.**

Cell-associated IgG binding activities were measured in both seropositive and seronegative vaccinees using 1:6250 dilution of sera (month 0 and month 7) incubated with Towne gB-transfected 293T cells. p values from Wilcoxon matched-pairs signed rank test are reported.  *p<0.05, **p<0.01, ***p<0.001.

**Supplementary Figure 6. Comparison of HCMV-specific neutralizing antibody response among mRNA-1647 seronegative vaccinees and naturally infected individuals.**
Epithelial neutralization antibody responses against AD169r (A) in ARPE-19 cells and fibroblast neutralizing antibody responses in HFF-1 cells against AD169r (B) and Towne (C) was compared between mRNA-1647 seronegative vaccinees at month 7 and infected individuals.

Open brown/light green/green circle denotes the seropositive samples at baseline from the mRNA-1647 vaccine cohort while open purple circle denotes the in-house HCMV seropositive subjects in some assays to increase the statistical power. But the statistical analysis indicated the increased sample # didn’t change the conclusion.. P values from non-parametric t-tests are reported (Mann-Whitney). *p<0.05, **p<0.01, ***p<0.001.

**Supplementary Figure 7. gB AD-specific IgG response mapping in the mRNA-1647 vaccine cohort.**
The antigenic domain (AD) 1 (A), AD4 (B), AD5 (C), and AD4&5 (D) were mapped via BAMA, and AD2S1 (E) was measured by ELISA before vaccination (month 0), and at the peak response timepoint (month 7) for both seropositive and seronegative vaccinees in mRNA-1647 vaccine cohort. The dotted line indicated as the cut-off for positivity (MFI=100). p values from Wilcoxon matched-pairs signed rank test are reported.  *p<0.05, **p<0.01, ***p<0.001.

**Supplementary Figure 8. Normalized vaccine antigen-specific IgG FcR binding corroborated that mRNA-1647 induced higher PC-specific FcR-binding IgG antibody responses than gB-specific FcR-binding IgG responses to the same formulation.**
The normalized FcγR1A (A), FcγR2A (B), FcγR2B (C), FcγR3A (D), and FcRN (E) IgG antibody responses were compared among gB and PC antigen components in the same vaccine formulation in seronegative vaccinees of the mRNA-1647 vaccine cohort. HCMV antigen-specific IgG binding to FcRs was normalized as a ratio of total IgG binding measured by ELISA. p values from non-parametric t-tests are reported (Mann-Whitney). *p<0.05, **p<0.01, ***p<0.001.

**Supplementary Figure 9. Comparison of gB AD-specific IgG mapping between mRNA-1647 and gB/MF59 vaccine cohorts.**

The gB AD1 (A), AD4 (B), AD5 (C), AD4&5 (D) and AD2S1 (E) responses were compared between 2 vaccine cohorts at peak responses (month 7) upon vaccination. P values from non-parametric t tests are shown (Mann-Whitney). *p<0.05, **p<0.01, ***p<0.001.

**Supplementary Figure 10. Comparison of normalized gB-specific FcR-binding IgG responses among the two vaccine cohorts.**
The normalized gB-specific FcgR1A (A), FcgR2A (B), FcgR2B (C), FcgR3A (D), and FcRN (E) IgG antibody responses were compared among mRNA-1647 and gB/MF59 vaccine cohort. HCMV gB-specific IgG binding to FcRs was normalized as a ratio of total IgG binding measured by ELISA. Additionally, the HCMV infected individuals (S+) plotting as blue circle from seropositive vaccinees at baseline from the mRNA-1647 vaccine cohort were also included in the comparison. p values from non-parametric t-tests are reported (Mann-Whitney). *p<0.05, **p<0.01, ***p<0.001.

**Supplementary Figure 11. Amino acid sequence alignment of subunit component of PC proteins between 2 related human CMV strains.**

The amino acid (AA) sequences of subunit gH, gL, UL128, UL130, and UL131A proteins from the HCMV Merlin strain to form the PC immunogen component in the mRNA-1647 vaccine were aligned with those from the VR1814 strain used to evaluate the PC-specific antibody responses in various assays in this report. AA numbering is according to Merlin reference strain. gH, gL, UL128, UL130, and UL131A from the VR1814 shared 95.96% (30aa), 98.2% (5aa), 99.41% (1aa), 98.6% (3aa), and 100% identity with those from Merlin strain.

**Supplementary Figure** **12. Amino acid sequence alignment of gB from different human CMV virus strains.**
The full-length gB amino acid (AA) sequences were aligned from HCMV strains Merlin, Towne, TB40/E, AD169, and VR1814, gB proteins or viruses from which strains were used for various serological assays for humoral response evaluations in this report. Among them, the full-length Merlin gB was used to encode the gB vaccine immunogen component for the mRNA-1647 vaccine, while the Towne gB with transmembrane region truncated and furin cleavage site mutation was used to encode the vaccine immunogen in gB/MF59 vaccine. AA numbering is according to the reference strain Merlin. The Towne, TB40/E, AD169, and VR1814 shared 99.34% (6aa), 99.56% (4aa), 95.82% (47aa), 92.62% (67aa) AA identity in gB protein with the Merlin strain. The gB genotype for each strain is listed in the parenthesis next to the strain according to the reference (21).

**Supplementary Figure 13. Images of HCMV neutralization in ARPE-19 cells against AD169 virus.**

The images illustrated one representative image file from cell control (left), virus control (middle), and irrelevant negative control (Synagis: right) during performing the neutralization assay against AD169r in ARPE-19 cells (A). The serial image files show the different levels of infected ARPE-19 cells (green) to exhibit the neutralization potency by various concentrations of positive control (B). In addition, the graph demonstrated the neutralization curve for positive control to determine the IC50 value (1.312 ng/uL) against the AD169r virus in ARPE-19 cells (C), mirroring panel B. The bold dotted line denoted the max percentage of infected cells while the thin dotted line indicated the 50% of infected cells to determine the IC50. Similarly, the serial raw image files to show the neutralization potency for different sera dilutions from one representative seronegative sample (D, SN-03) and one seropositive sample (E, SP-04) were displayed at both month 0 and peak responses at month 7 upon vaccination with mRNA-1647, respectively. The green cells indicated AD169r infected ARPE-19 cells after the cells were stained with anti-HCMV IE-1 antibody as 1^st^ antibody and anti-mouse IgG-AF488 as 2^nd^ antibody then the image data were acquired. DAPI stained the cell nuclei to quantify the total cells. Both IC50 and ID50 was determined using non-linear regression analysis (Sigmoidal, 4PL) using GraphPad Prism. The numbers in red on panels B, D-E indicate the ID_50_ cut-off.

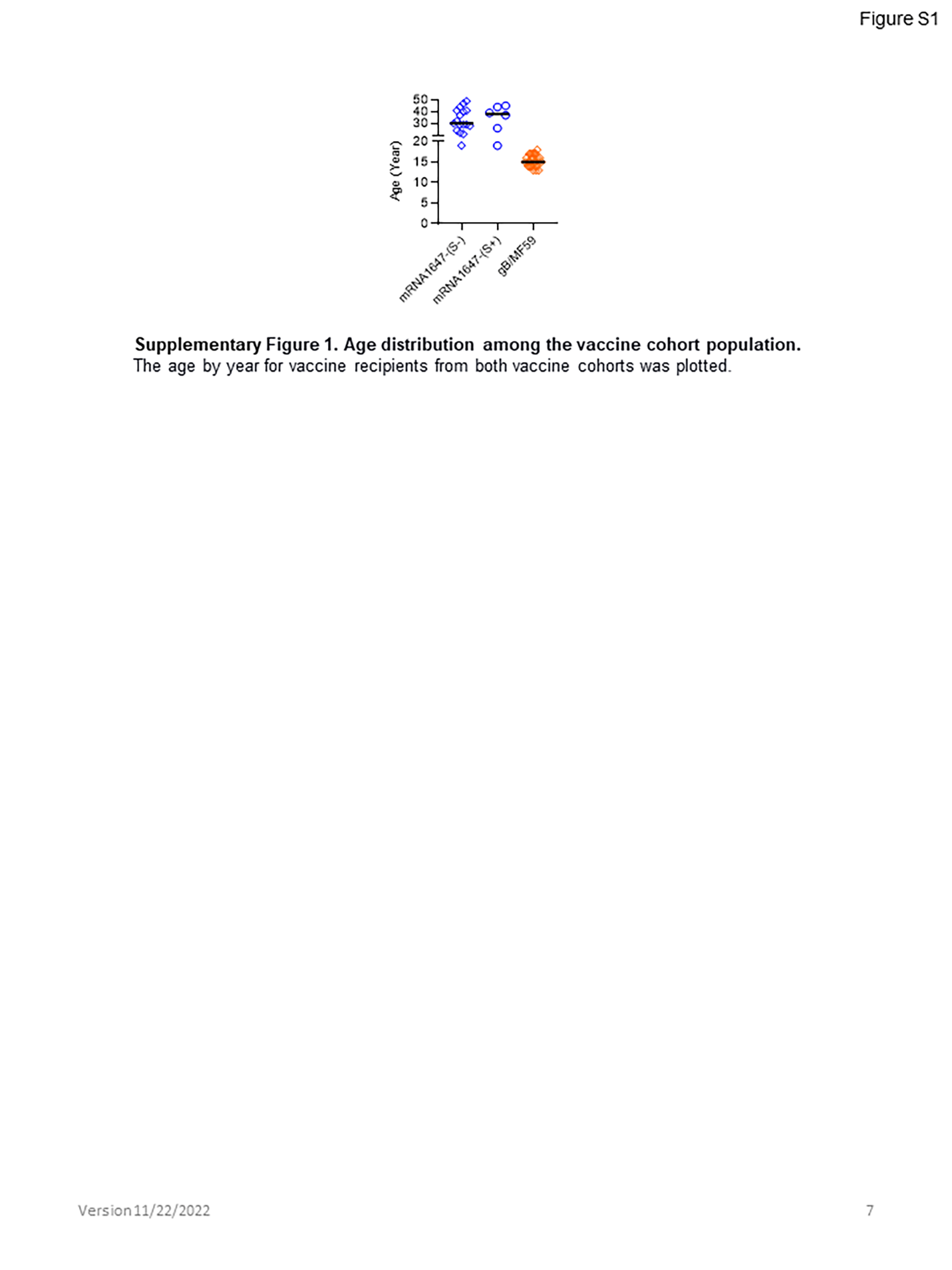

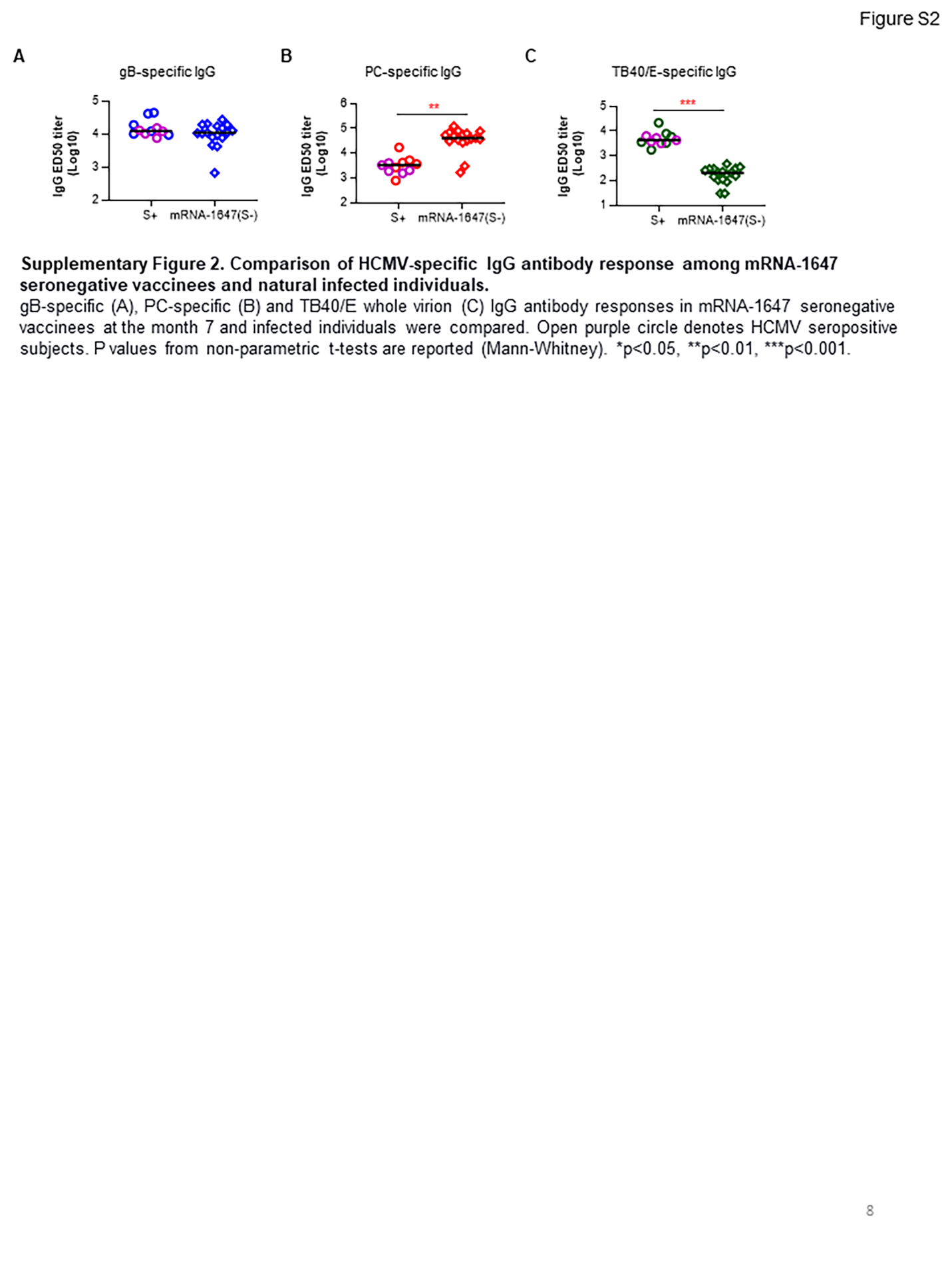

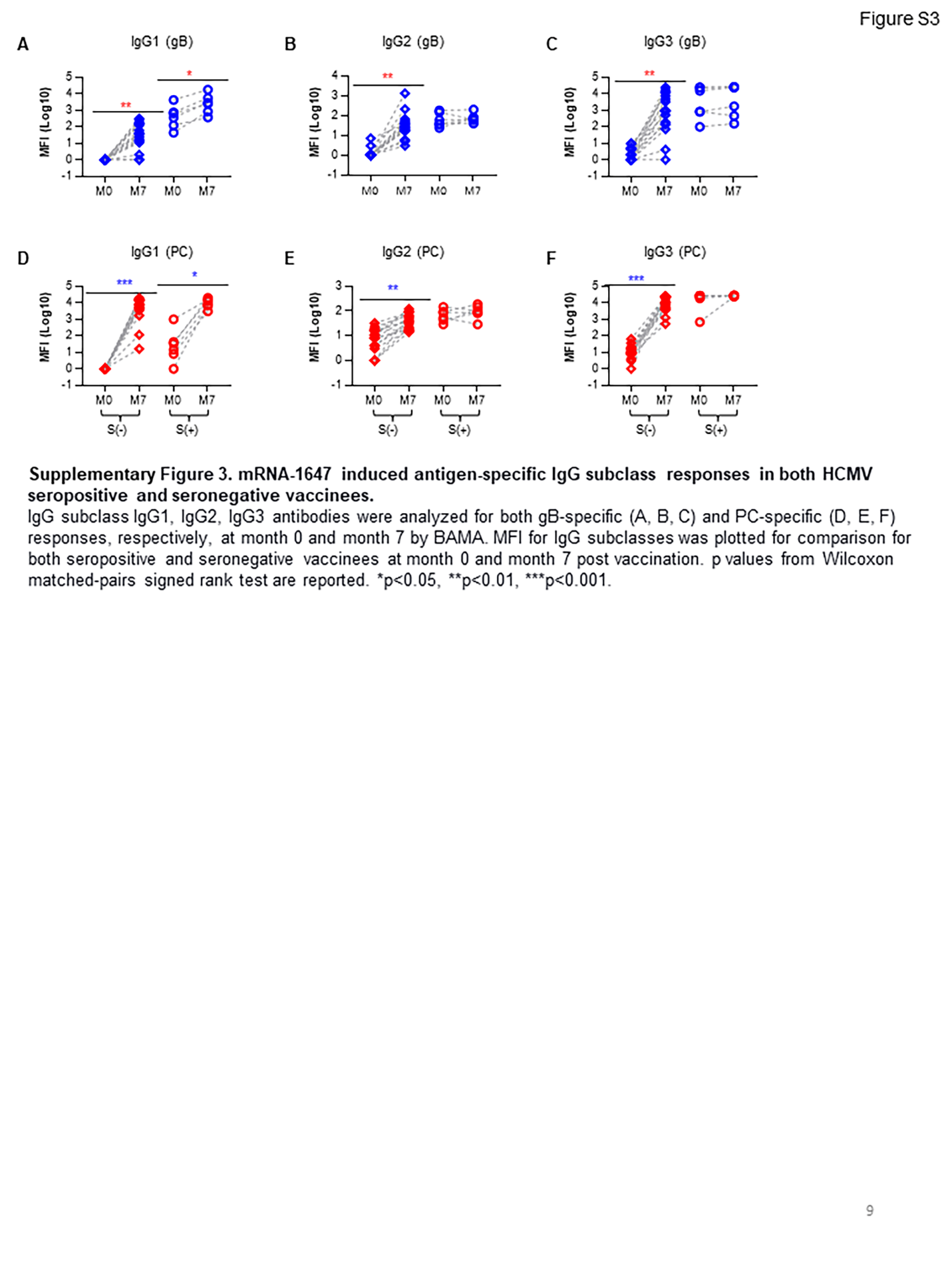

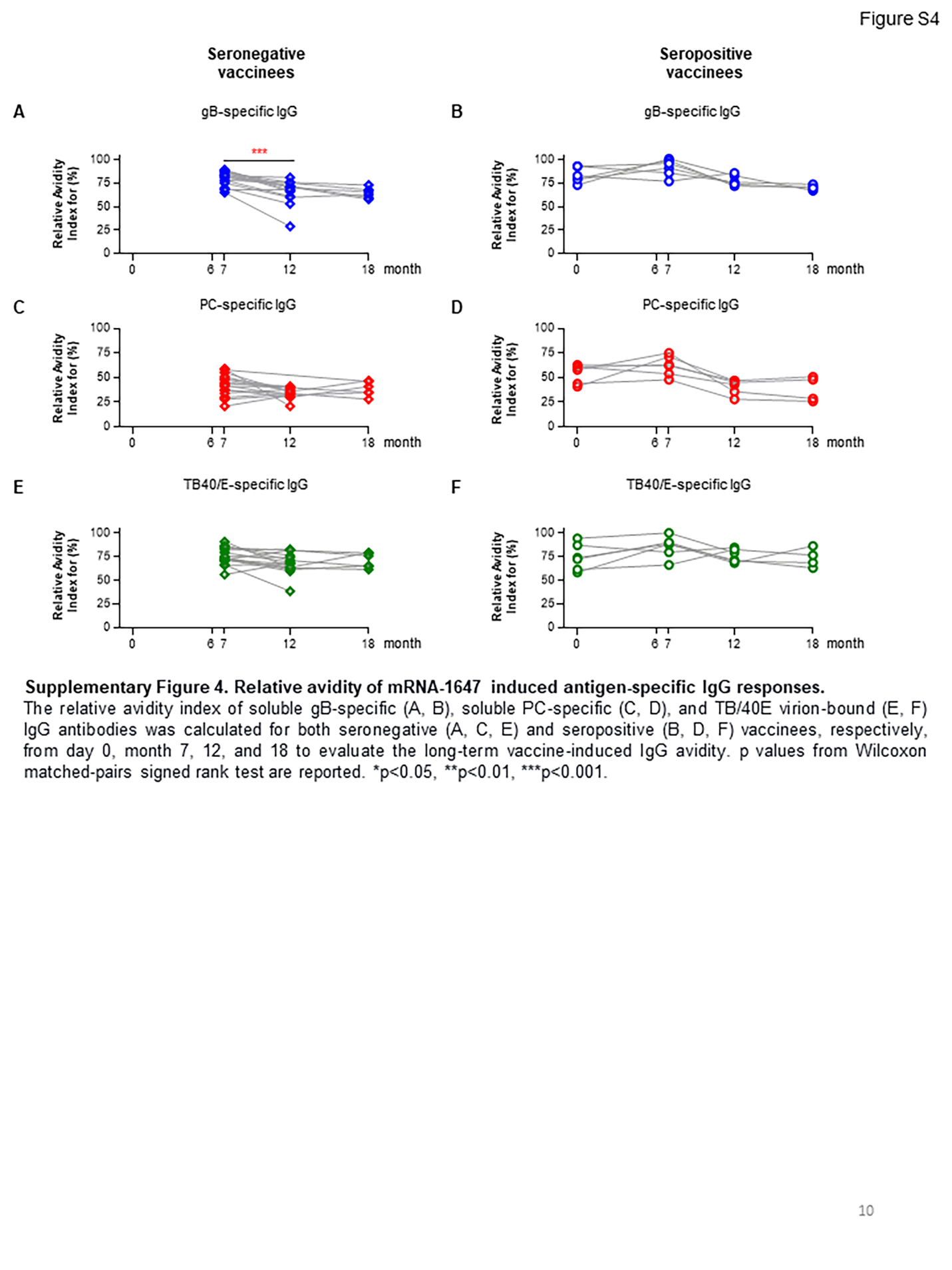

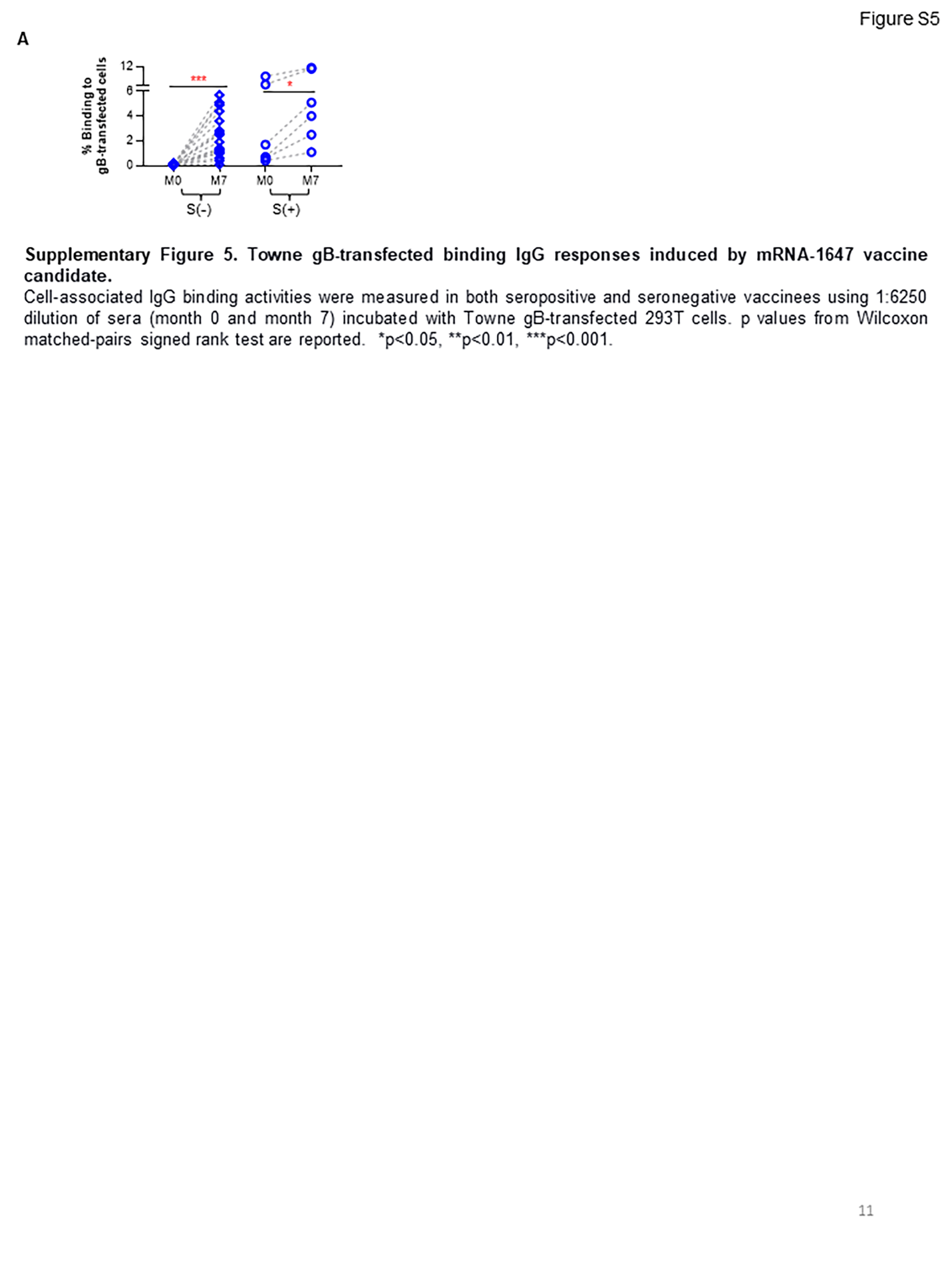

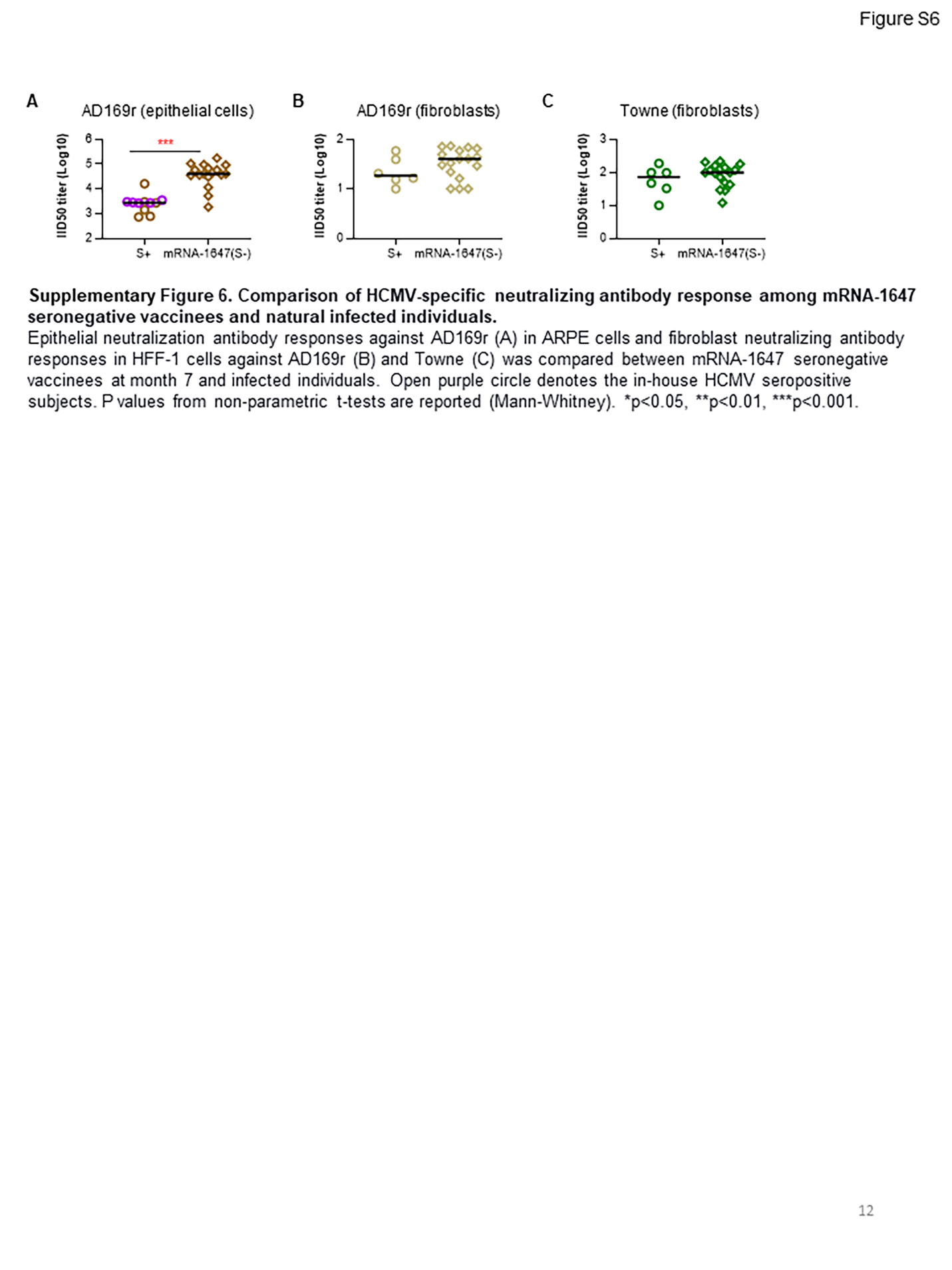

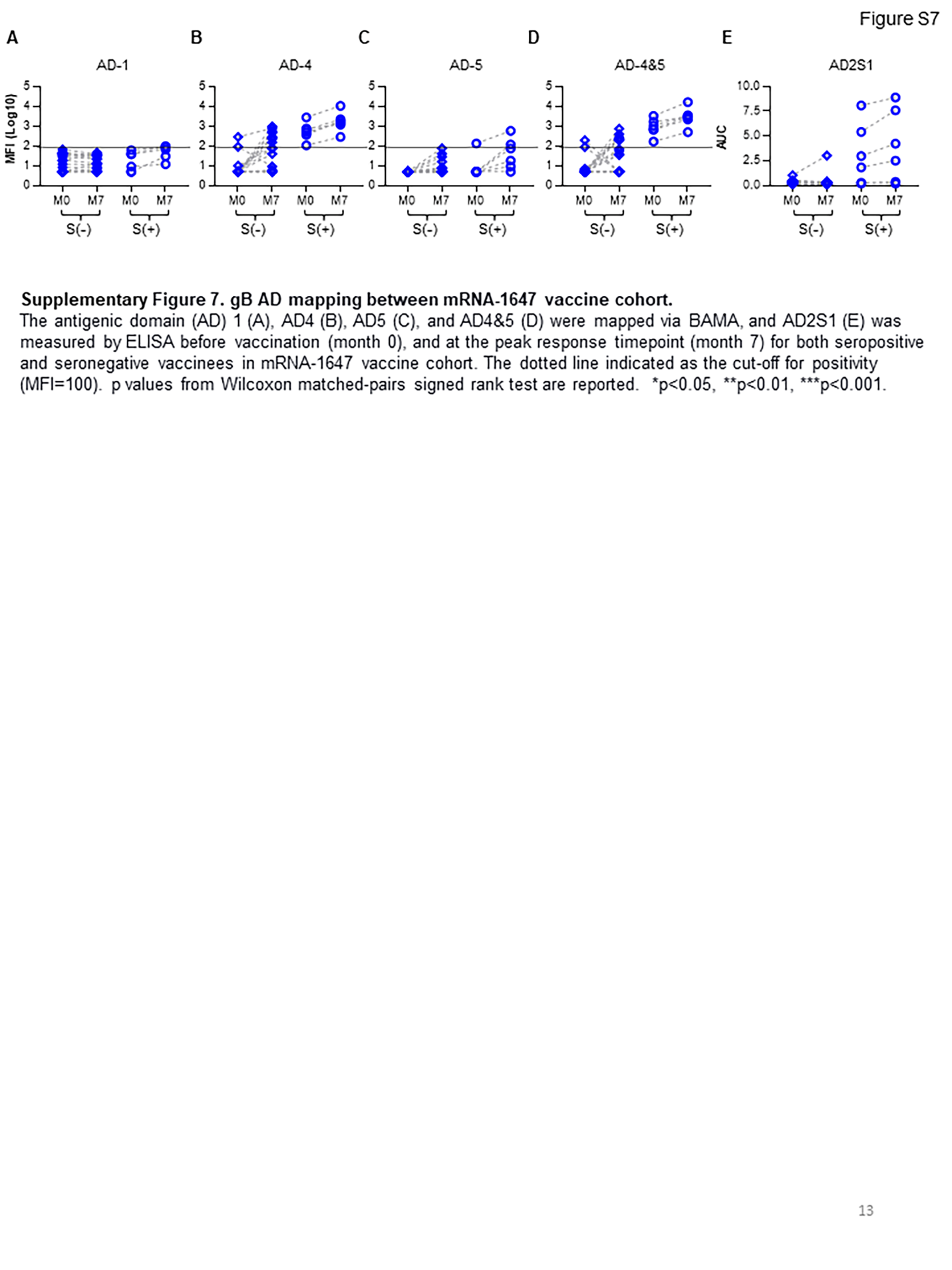

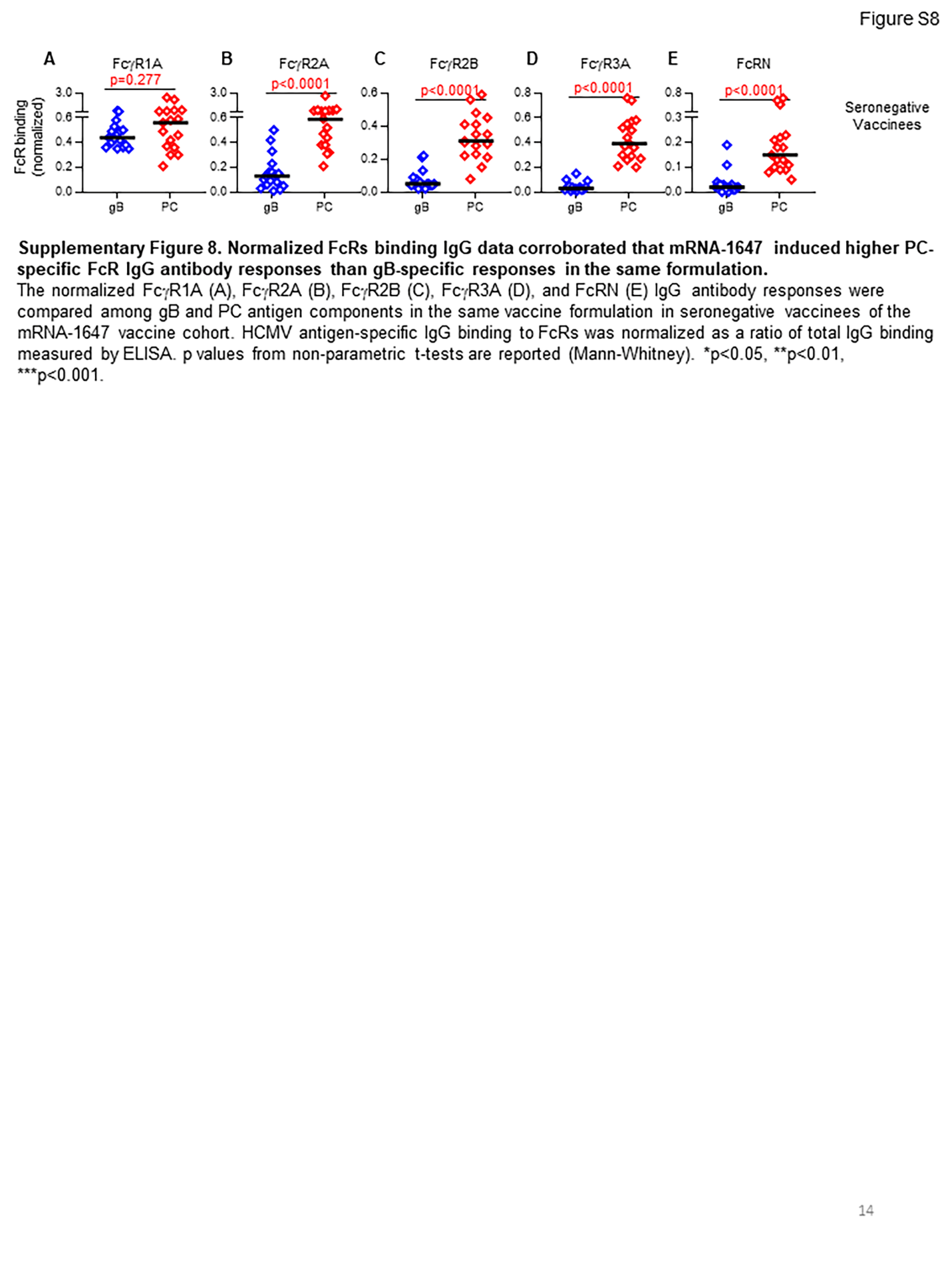

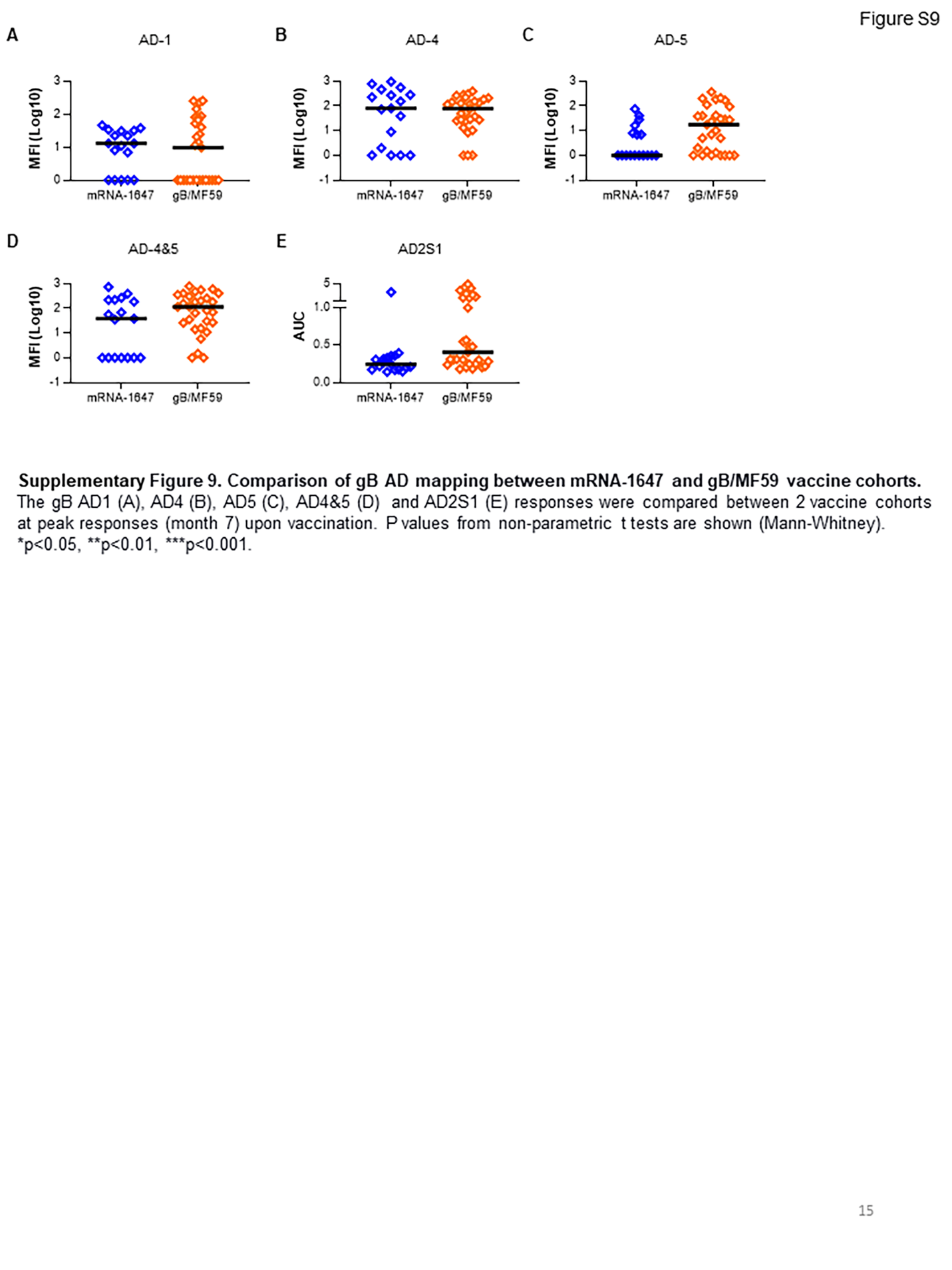

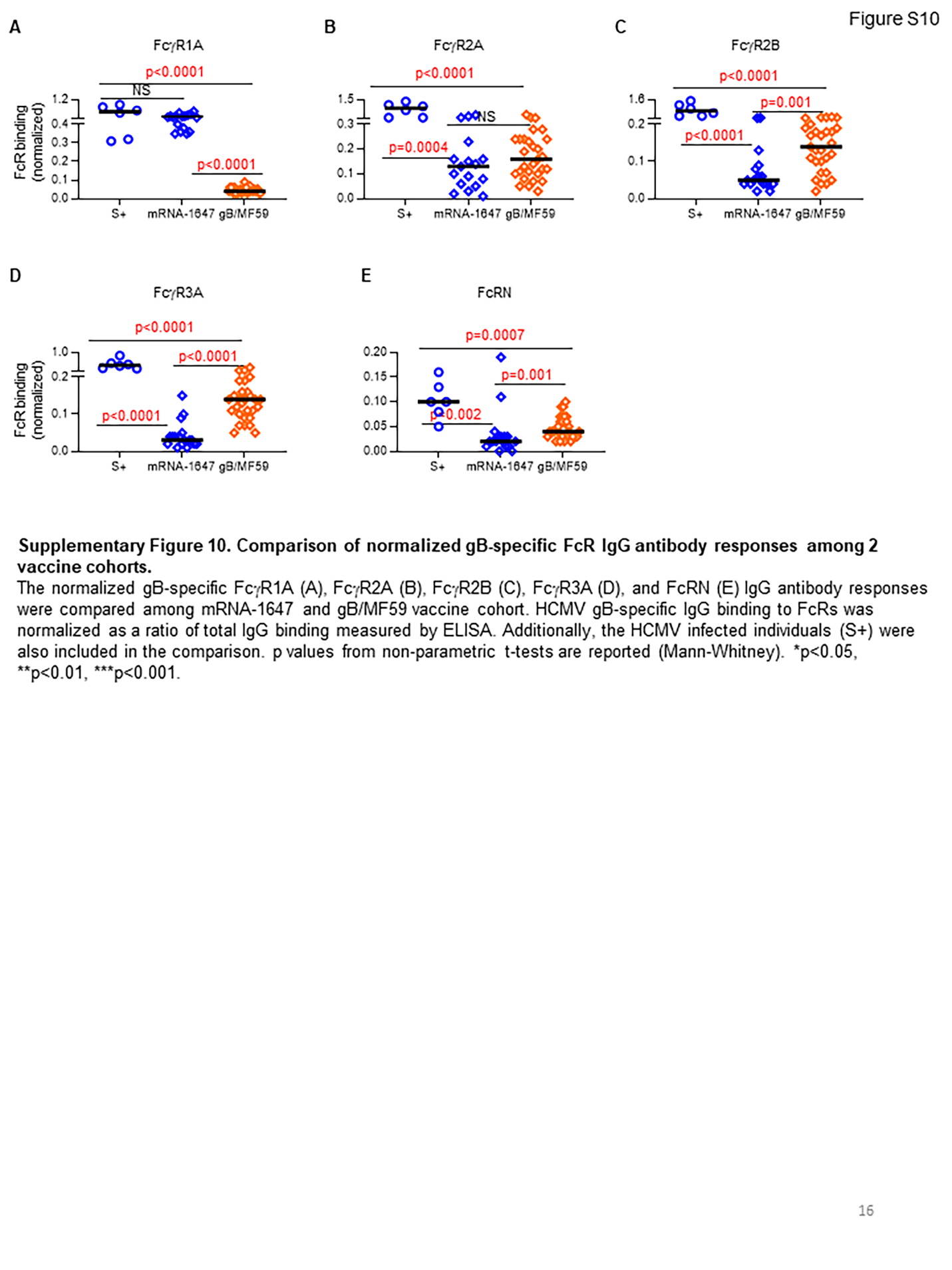

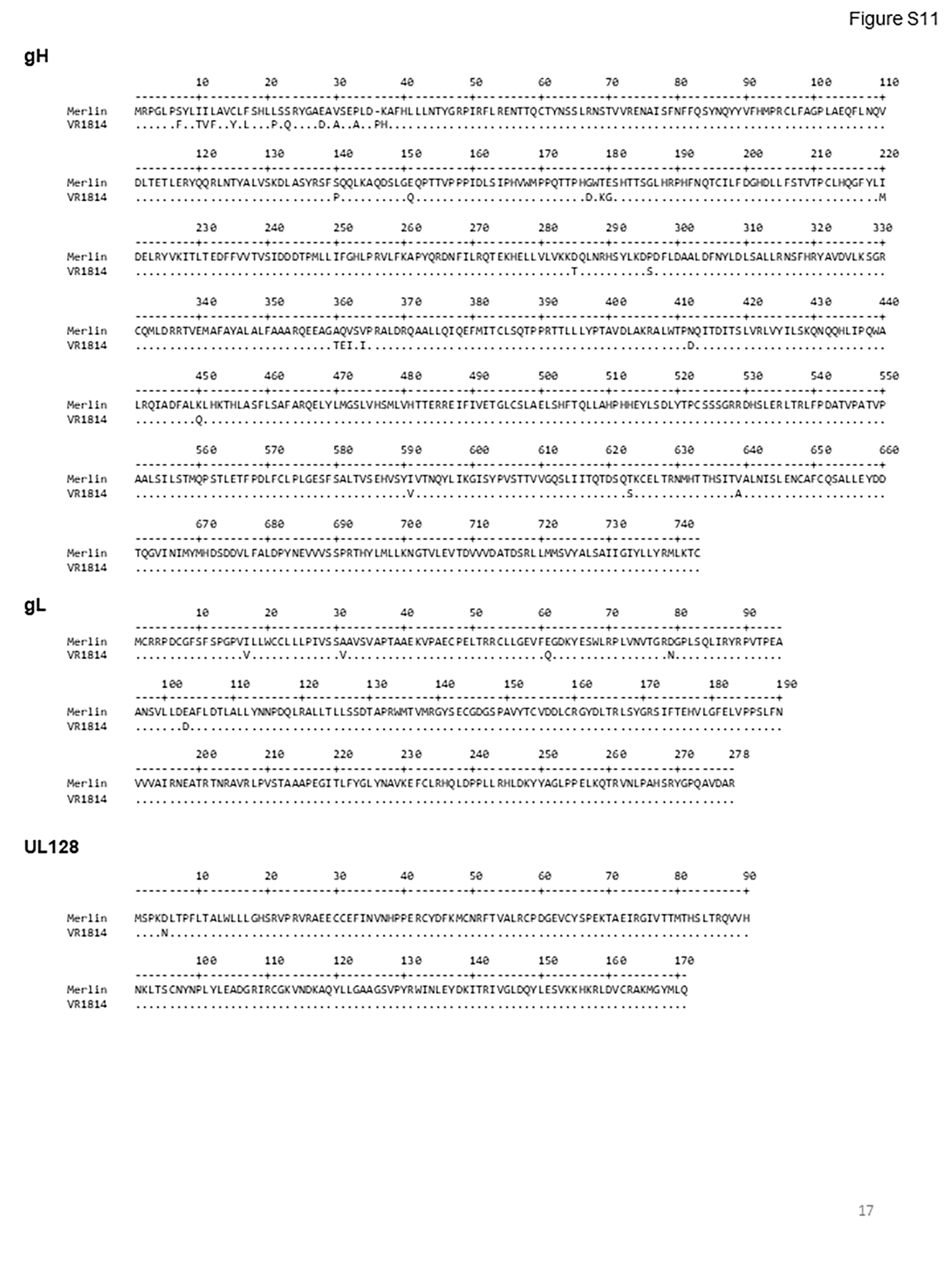

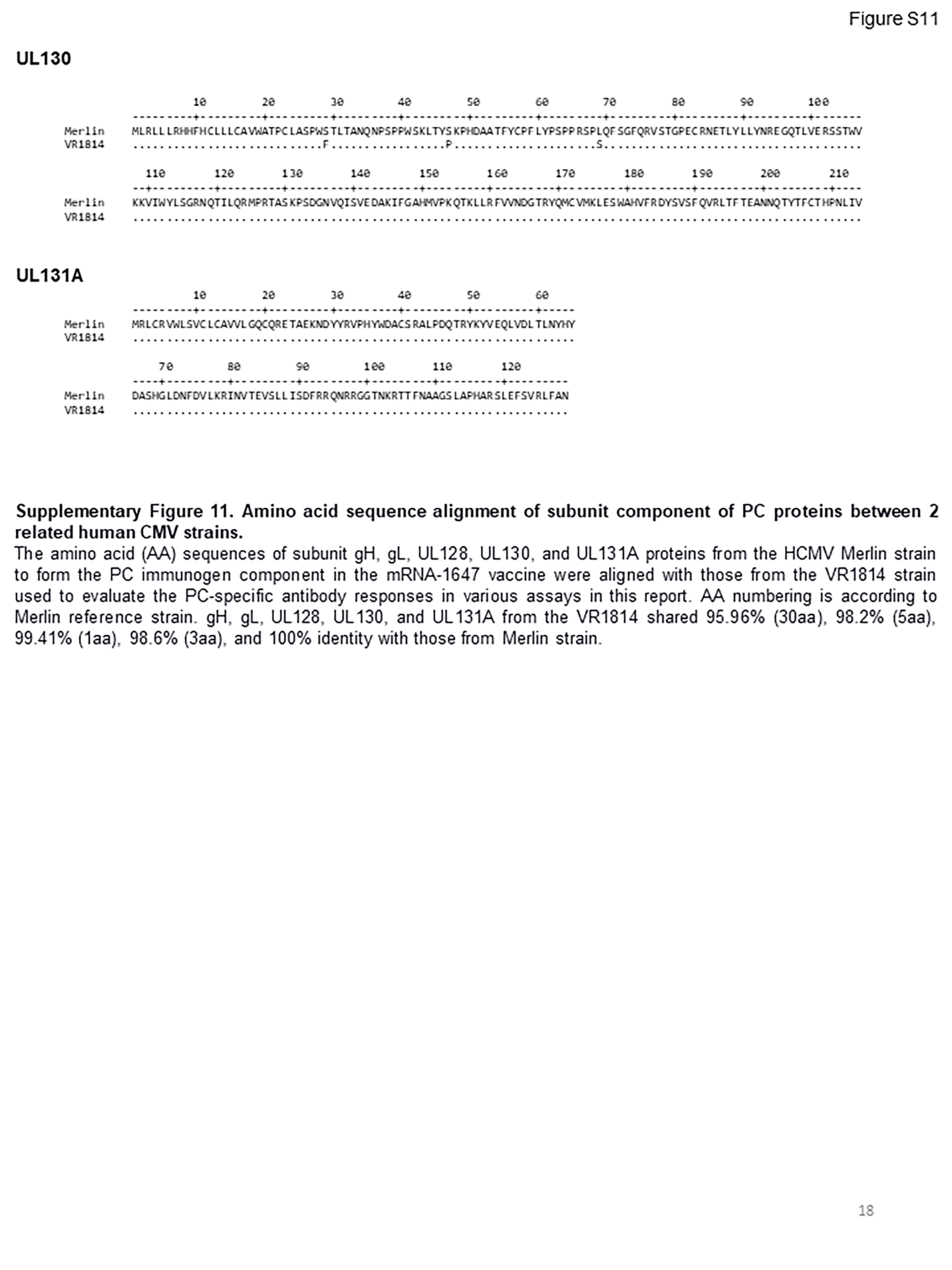

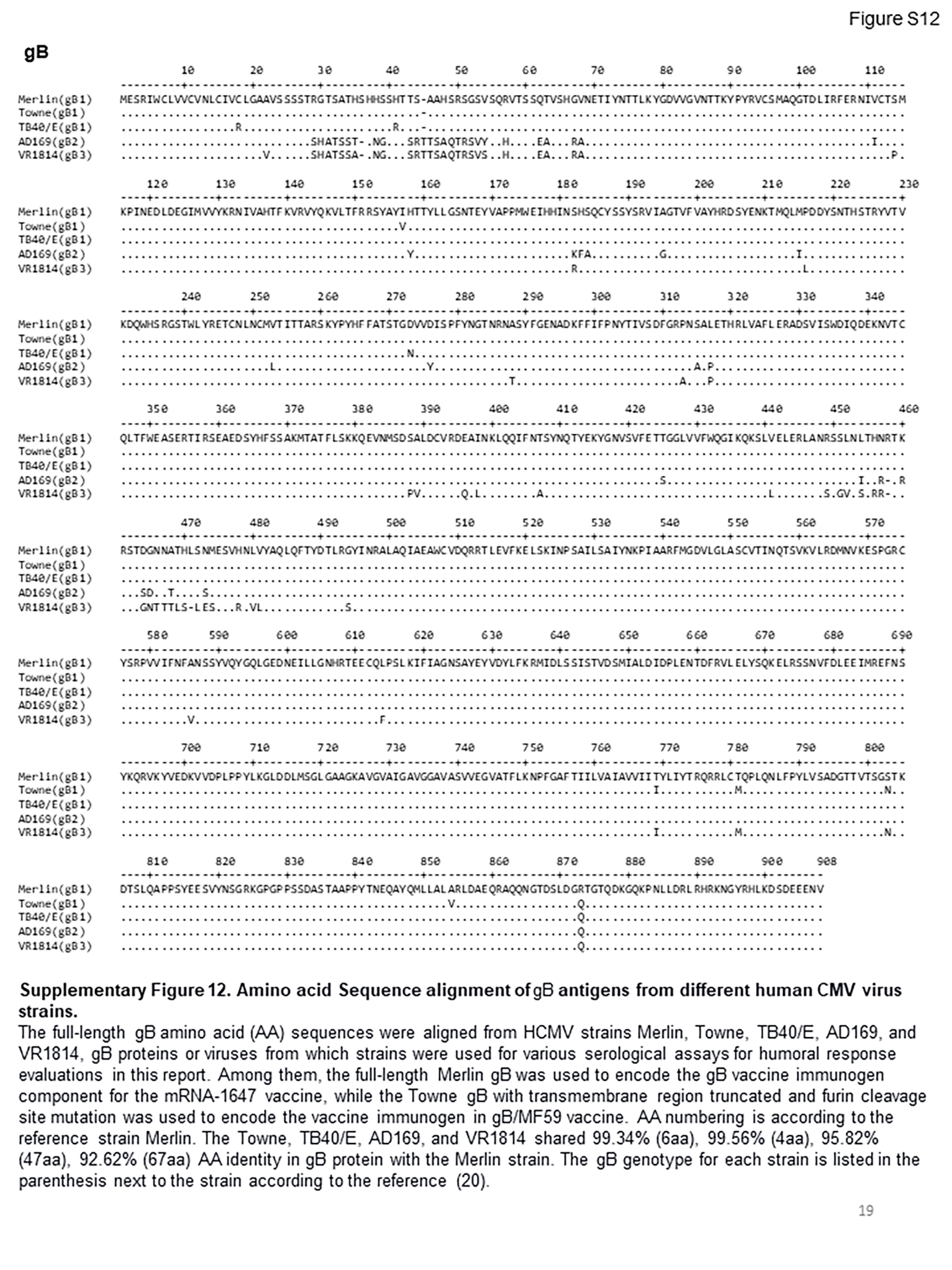

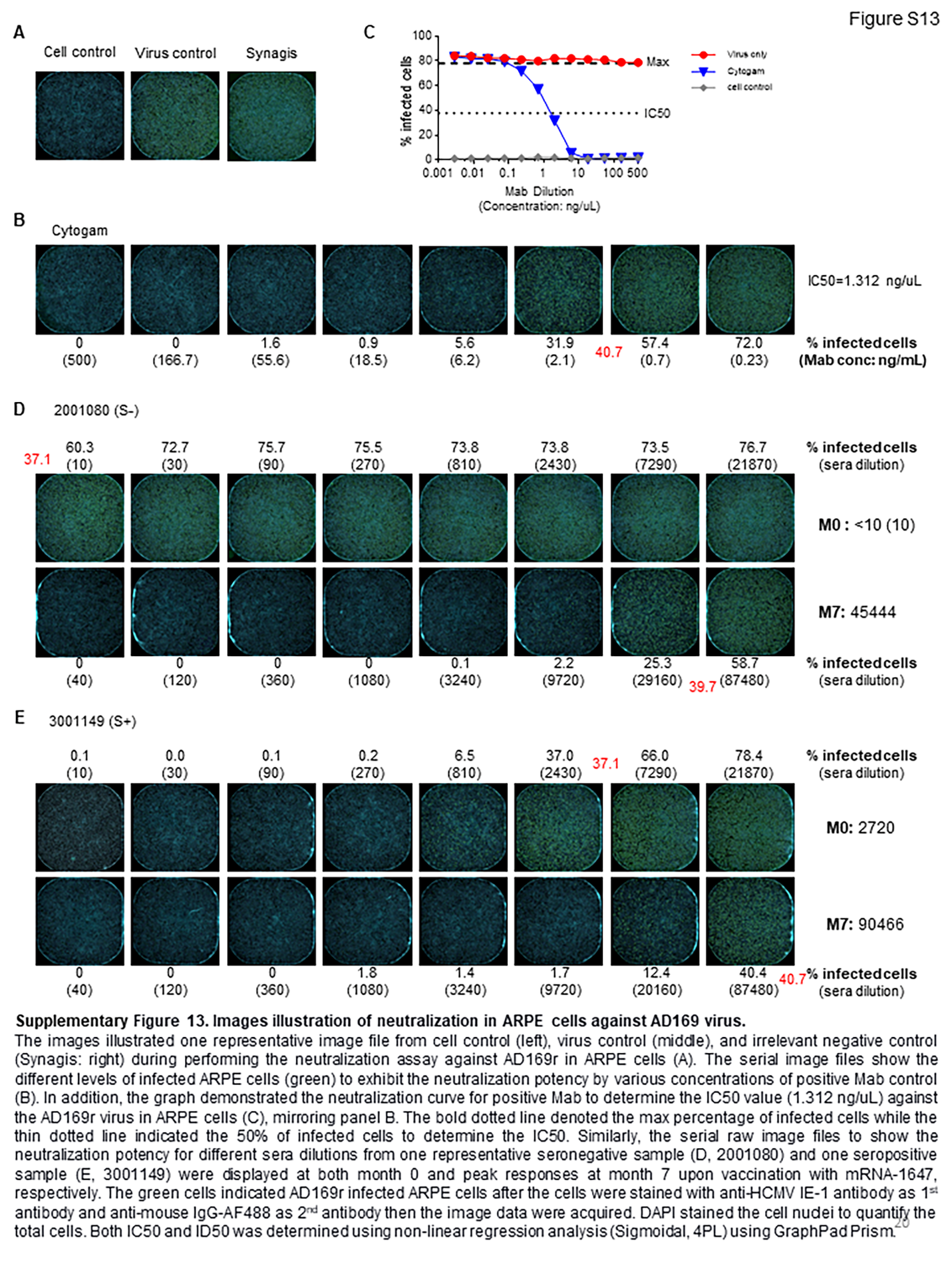
